# Similar discrimination, different behaviour under synthetic occlusion: confidence and attribution reproducibility of deep learning models for intraoral ultrasonography

**DOI:** 10.64898/2026.11.03.26367069

**Authors:** Goun Jung, Yu-Ran Heo, Kyung-Seok Hu, Hee-Jin Kim

**Affiliations:** Division of Anatomy and Developmental Biology, Department of Oral Biology, Human Identification Research Institute, BK21 FOUR Project, Yonsei University College of Dentistry, Seoul 03722, Republic of Korea

**Author notes:** Corresponding author: Hee-Jin Kim, Division of Anatomy and Developmental Biology, Department of Oral Biology, Yonsei University College of Dentistry, 50-1 Yonsei-ro, Seodaemun-gu, Seoul 03722, Republic of Korea.

**Keywords:** intraoral ultrasonography, buccal mucosa, deep learning, occlusion testing, explainable artificial intelligence

## Abstract

**Objectives:** To determine whether similar baseline discrimination conceals different responses to synthetic occlusion, whether loss of discrimination is accompanied by reduced confidence, and whether spatial attributions are reproducible in intraoral ultrasonography models.

**Methods:** We analysed 477 images from 40 healthy volunteers, using acquisition position to label superior and inferior subregions. Five architectures were evaluated across five training seeds on a subject-independent test set of 97 images from eight volunteers. Images underwent constant-intensity occlusion at boundaries spanning 30– 70% of image depth and an area-matched band-occlusion control. Attribution analyses assessed variation across seeds and class-activation methods and compared attribution-guided insertion with random insertion.

**Results:** Seed-mean areas under the receiver operating characteristic curve ranged from 0.958 to 0.972, with rankings reversing across seeds. EfficientNet-B0 fell below 0.60 in 49 of 50 boundary-sweep conditions, compared with zero to two for the other architectures; the difference persisted with area-matched masking. Under 50% occlusion, its mean confidence on incorrect predictions rose to 0.989 and expected calibration error from 0.094 to 0.478. Between two inference runs, EfficientNet-B0’s per-seed occluded AUCs differed by up to 0.161; seed means were 0.51–0.55 in both. Attribution patterns varied across seeds and methods.

**Conclusions:** Similar baseline discrimination concealed different occlusion responses, and loss of discrimination could coexist with high confidence. Attribution reproducibility provided a further constraint on interpretation. These findings concern the evaluated pipelines and synthetic masks; they do not establish anatomical mechanisms or clinical reliability.

**Advances in knowledge:** Joint evaluation of perturbation sensitivity, confidence under perturbation and attribution reproducibility distinguished model behaviours that baseline discrimination alone did not reveal in intraoral ultrasonography.

## 1. Introduction

Intraoral ultrasonography provides real-time imaging without ionizing radiation. It has been investigated for ultrasound-guided resection of buccal squamous cell carcinoma [1] and for preoperative measurement of tumour depth and surgical margins [2,3]. Other applications include oral and maxillofacial assessment [4], mucosal characterization [5], salivary gland assessment [6] and imaging with dedicated intraoral probes [7]. Interpretation depends on probe placement and recognition of the imaged tissue layers. Deep learning has been applied to dental implant planning in cone-beam computed tomography [8], periapical disease detection in dental radiographs [9] and ultrasound in other specialties [10,11]. Its use in intraoral ultrasonography remains comparatively limited, motivating evaluation of model behaviour beyond classification performance.

The buccal mucosa is a useful setting for this methodological evaluation. It provides tissue for buccinator musculomucosal flaps [12] and mucosal grafts [13]. Ultrasonographic descriptions of facial tissue layers provide a general anatomical framework for interpreting such images [14]. In the present study, the superior and inferior acquisition regions were defined by probe position relative to the underlying musculature (Section 2.2). Differences in the imaged tissue layers motivated depth-oriented perturbations, without assuming that a particular tissue boundary was the only discriminative feature.

For models with similar classification performance, a practical question is whether they also behave similarly when image information is altered. Baseline discrimination does not characterize dependence on image features or responses to altered inputs [15,16]. Confidence and visual attribution offer additional information, but their usefulness depends on how they behave when performance deteriorates and on whether the explanations are reproducible. A Grad-CAM overlay [17] from one trained model may appear anatomically plausible without establishing either property [18–22]. This leaves a model-selection and interpretation problem beyond the established observation that accuracy alone is insufficient: performance, confidence and attribution may give different accounts of the same model.

We chose superior–inferior buccal subregion classification as a methodological test case. Because the operator knows the probe position, the labels are fixed at acquisition and independent of retrospective image interpretation, and the expected anatomical differences motivate depth-oriented perturbations. For the same reason, the task is not intended for immediate clinical use; it offers a controlled setting in which to establish evaluation practices that can be applied to future clinical intraoral ultrasound models. The labels do not, however, identify which image features distinguish the classes or provide a pixel-level anatomical reference. The task permits comparison of model responses to controlled image changes without treating those changes as definitive tests of anatomical understanding.

We evaluated five architectures spanning residual, dense and compound-scaled convolutional families, a modernized convolutional design (ConvNeXt-Tiny) and a vision transformer (DeiT-Tiny), each across five random seeds. We asked whether similar baseline discrimination conceals different responses to partial image occlusion; whether deterioration in discrimination is accompanied by reduced confidence or instead by confident errors; and whether attribution patterns remain reproducible across training seeds and class-activation methods. Boundary sweeps and an area-matched control examined the scope of the occlusion response, while comparison with random insertion assessed the support for attribution-based interpretation. These questions concern the evaluated training pipelines and controlled perturbations, rather than intrinsic properties of architecture families or demonstrated clinical failure.

## 2. Materials and Methods

### 2.1 Study population and data acquisition

This study was approved by the Institutional Review Board of Yonsei University Dental Hospital (approval no. 2-2023-0050; approved on 19 October 2023), covering both the original anatomical investigation and the secondary deep learning analysis presented herein, and was conducted in accordance with the Declaration of Helsinki. The final cohort comprised 40 healthy volunteers (11 male, 29 female), mean age 33.4 ± 12.0 years (range 20–60) at acquisition. All provided written informed consent before enrolment. The images were originally acquired for anatomical investigation of the buccal mucosa under routine clinical conditions, not under a protocol prospectively designed for deep learning. A high-frequency intraoral linear transducer was used for both superior and inferior buccal subregions within a single session per subject; every subject contributed to both classes, matching subject-level acquisition parameters between them. Of the 531 images acquired, 54 (10.2%) were excluded at quality assessment for file corruption, inadequate anatomical visualization, or colour Doppler overlay (Supplementary Methods S2). The final analytic dataset comprised 477 images (superior: 240; inferior: 237). Volunteers were recruited by public notice and imaged in July and August 2024; the parent study’s sample size was determined a priori, whereas no separate sample size was prespecified for this secondary analysis, which used every image that passed quality assessment (Supplementary Methods S11). Equipment and parameter distributions are given in Supplementary Methods S1 (Supplementary Tables S1 and S3).

Measured acquisition parameters were compared between classes across the dataset, within subjects and within the test set (Supplementary Methods S3, Table S2). These exploratory comparisons detected no association (all P ≥ 0.750), but non-significance does not establish independence or exclude unmeasured acquisition confounding.

### 2.2 Anatomical labelling and data partitioning

Class labels were determined at the time of acquisition by the scanning position selected by the operating anatomist, who holds a doctorate in human anatomy and more than five years of experience in intraoral ultrasonography of the buccal mucosa. The superior class was defined as the upper buccal subregion overlying the zygomaticus major and levator anguli oris musculature; the inferior class was defined as the lower buccal subregion overlying the buccinator muscle belly. General ultrasonographic descriptions of the facial musculature and tissue layers provide the anatomical context for these acquisition positions [14]. In images acquired at the inferior position, a hyperechoic dermis line was typically visible in the far field. Defining labels by scanning position rather than post hoc image reading anchored class assignment to landmarks identified during scanning and reduced reliance on retrospective image interpretation.

The final dataset showed near-perfect class balance (superior: n = 240, 50.3%; inferior: n = 237, 49.7%; difference < 1%). Data were partitioned at the subject level into training/validation (32 subjects, 380 images) and held-out test (8 subjects, 97 images; superior 50.5%, inferior 49.5%) sets with no subject overlap, to prevent leakage [23]. Split records were saved before training.

### 2.3 Preprocessing

Prior to dataset construction, each ultrasound frame was cropped to retain only the rectangular B-mode imaging sector. All peripheral display elements — vendor identifiers, patient information, gain, depth and frequency annotations and timestamps — were excluded, so that the model input consisted exclusively of acoustic image content. The cropped images were resized to 224 × 224 pixels and converted into three-channel greyscale. Pixel values were normalized using the mean (0.316) and standard deviation (0.171) computed exclusively from the training set to prevent data leakage. The vertical image axis represents tissue depth, from the near-field buccal mucosa toward the far-field skin. Geometric augmentation, including flipping and rotation, was not used, so that the depth orientation of the images was preserved.

### 2.4 Deep learning architectures

Six architectures were trained: ResNet-50 [24], DenseNet-121 [25], EfficientNet-B0 and EfficientNet-B7 [26], ConvNeXt-Tiny [27] and DeiT-Tiny [28]. EfficientNet-B7 was excluded after cross-validation and before any test evaluation (Section 3.3), leaving the five architectures reported throughout. All the models were initialized with ImageNet pre-trained weights [29], and the final classification layers were replaced with two-class output layers. They span distinct design principles: additive residual propagation (ResNet-50), dense connectivity (DenseNet-121), compound scaling with depthwise-separable convolutions and squeeze-and-excitation blocks (EfficientNet [26]), a modernized convolutional design (ConvNeXt-Tiny) and a vision transformer (DeiT-Tiny). EfficientNet-B0 (4.0M parameters) and EfficientNet-B7 (63.8M) span a wide capacity range; ConvNeXt-Tiny and DeiT-Tiny test whether the findings generalize beyond residual and dense convolutional designs. All five retained architectures were evaluated across five random seeds; three were additionally carried through the detailed best-fold analyses (Supplementary Table S10). In every architecture all layers were fine-tuned; no backbone layers were frozen, and the optimizer was applied to the full parameter set. ImageNet pre-training was used to mitigate the constraints of small-sample medical image training [30]. All pipelines were implemented in Python using PyTorch on Google Colab.

### 2.5 Hyperparameter optimization

Hyperparameters were optimized with Optuna v4.7.0 [31] using Tree-Structured Parzen Estimator sampling, with 20 trials per architecture. For ResNet-50, DenseNet-121 and EfficientNet, Fold 1 was the tuning set; the search covered learning rates of 1 × 10⁻⁵ to 1 × 10⁻⁴ (log scale), batch sizes of 8, 16 or 32, Adam or SGD, and 20–50 maximum epochs. ConvNeXt-Tiny and DeiT-Tiny were tuned and trained on the saved Fold 2 partition. ConvNeXt-Tiny used the same learning-rate range with Adam or AdamW; DeiT-Tiny used 1 × 10⁻⁶ to 5 × 10⁻⁵ with AdamW. Batch-size and epoch ranges were unchanged; AdamW used weight decay 0.05. Early stopping used validation F1 with patience 5. These architecture-specific settings and single-fold tuning may affect comparisons [32]. Supplementary Methods S4 and Table S4 report the settings.

### 2.6 Performance evaluation

Five-fold subject-independent cross-validation with GroupKFold [33] was performed for ResNet-50, DenseNet-121 and EfficientNet (approximately 6–7 validation subjects per fold). For the detailed single-run analysis, the fold model with the highest validation F1 was evaluated on the held-out test set. The five-seed experiments used the saved Fold 2 training/validation partition; ConvNeXt-Tiny and DeiT-Tiny were evaluated across seeds on this partition rather than by a separate five-fold cross-validation. Seed averages therefore quantify training-run variation on a fixed partition, not variation across data splits. Held-out metrics were precision, recall, F1 and the area under the receiver operating characteristic curve (AUC). Inference time for the three best-fold models was measured over 10 repeated runs (batch size = 1, single GPU). Fold-level results are reported in Supplementary Table S5.

### 2.7 Explainability analysis

Grad-CAM [17] was used to visualize and quantify spatial attribution. Occlusion experiments then measured prediction sensitivity to removing specified image regions with a constant-intensity mask. These complementary analyses do not by themselves identify the anatomical features used by a model.

#### 2.7.1 Target-layer selection

For ResNet-50, DenseNet-121 and EfficientNet-B0, target layers were selected by comparing activation patterns across candidate layers (Supplementary Figure S1). Deeper layers gave higher mean activation but spatially concentrated, anatomically non-specific maps, whereas intermediate layers preserved spatial resolution; ResNet-50 (Layer3), DenseNet-121 (DenseBlock3) and EfficientNet-B0 (features[−3]) were used. ConvNeXt-Tiny used the final feature stage (model.features[−1]). DeiT-Tiny used the first layer normalization in the final transformer block (model.blocks[−1].norm1). For the non-distilled deit_tiny_patch16_224 model, the class token was removed and the remaining 196 patch tokens were reshaped to a 14 × 14 grid, then transposed to channel-first format before CAM calculation. The same target layers and token transformation were used for the alternative CAM methods; no layer-selection comparison is claimed for these two architectures.

#### 2.7.2 Quantitative Grad-CAM analysis

Three complementary spatial metrics were computed from Grad-CAM heatmaps for each test image: (1) mean activation intensity, the mean pixel value of the normalized heatmap; (2) the intensity-weighted vertical centroid (ȳ-norm), normalized to image height (0 = top, 1 = bottom); and (3) the upper-to-lower activation ratio (ULR), the ratio of mean activation in the upper image half to that in the lower, clipped at 10.0 to exclude instability from near-zero lower activation. Together they separate attention magnitude from spatial location. In addition, the faithfulness of Grad-CAM attributions was assessed via insertion and deletion analyses [34] with random-mask controls (the area under the curve for insertion and deletion as well as Average Drop). Average activation maps were generated by aggregating Grad-CAM heatmaps across all test images within each class to visualize global attention patterns.

#### 2.7.3 Representative case analysis

Correctly classified and misclassified cases were selected by classification confidence and examined for sonographic features that could account for the error.

### 2.8 Occlusion ablation experiment

To complement the visualization-based Grad-CAM analysis with a controlled test of perturbation sensitivity, an occlusion ablation experiment was performed on the trained models. For each test image (n = 97), two occluded variants were generated by replacing the upper or the lower region of the image with a constant value equal to the training-set mean intensity, corresponding to (1) upper-region occlusion and (2) lower-region occlusion. Occlusion was applied along the depth axis, because in ultrasound anatomical content is organized by depth; the boundary itself was a fixed geometric fraction of image height (50% in the primary analysis) and was not delineated anatomically. The retained and removed regions therefore differ systematically but not exactly in tissue content: near-field mucosal and superficial muscle layers lie predominantly in the upper region, and the far-field dermis echo predominantly, though not exclusively, in the lower region. We treat this correspondence as approximate and do not attribute any observed effect to a specific anatomical landmark.

Two inference runs of the same five-seed models are reported. The original run, performed at training time, produced Table 1. A follow-up run of the saved checkpoints in a different computing environment produced the boundary-sensitivity, area-matched, calibration and subject-cluster analyses (Section 2.9; Supplementary Methods S12). Between the two runs, per-seed AUCs differed by less than 0.002 at baseline for all architectures and by less than 0.003 for the other four architectures under occlusion, but by up to 0.161 for EfficientNet-B0 under occlusion. For each model and condition, class predictions, predicted probabilities, and standard performance metrics (accuracy, F1-score and AUC) were computed. Seed-averaged classification and occlusion performance are reported in Table 1, and the stability of Grad-CAM spatial metrics across seeds and three alternative CAM methods (Grad-CAM++ [35], Score-CAM [36], Layer-CAM [37]) in Supplementary Tables S7 and S8. Eigen-CAM and gradient-integration methods, which differ in formulation from class-activation mapping, were not evaluated.

**Table 1.** Classification and occlusion performance across five architectures and five random seeds. Held-out test AUC and F1 (seed mean ± SD), AUC under 50% upper- and lower-region occlusion (seed mean), mean AUC drop, and the number of seeds (of five) in which both occluded AUCs fell below 0.60. Values come from the original execution. SD describes training-seed variation, not subject-sampling uncertainty; follow-up estimates and subject-cluster intervals are in Supplementary Table S14.

| **Architecture** | **Test AUC** | **Test F1** | **Occluded AUC (upper)** | **Occluded AUC (lower)** | **AUC drop** | **Seeds collapsed** |
|---|---|---|---|---|---|---|
| ResNet-50 | 0.958 ± 0.026 | 0.888 ± 0.029 | 0.885 | 0.904 | 0.063 ± 0.021 | 0/5 |
| DenseNet-121 | 0.959 ± 0.020 | 0.906 ± 0.018 | 0.866 | 0.875 | 0.088 ± 0.019 | 0/5 |
| EfficientNet-B0 | 0.963 ± 0.013 | 0.903 ± 0.012 | 0.549 | 0.543 | 0.417 ± 0.044 | 5/5 |
| ConvNeXt-Tiny | 0.972 ± 0.008 | 0.908 ± 0.011 | 0.917 | 0.874 | 0.076 ± 0.013 | 0/5 |
| DeiT-Tiny | 0.965 ± 0.010 | 0.911 ± 0.030 | 0.851 | 0.861 | 0.109 ± 0.026 | 0/5 |

### 2.9 Occlusion boundary sensitivity, area-matched control and output diagnostics

The 50% occlusion boundary is an arbitrary geometric choice. Three additional analyses were therefore performed, all by inference only using the stored seed checkpoints; no model was retrained.

First, the boundary was varied. Each architecture, at each seed, was re-evaluated with the boundary at 30%, 40%, 50%, 60% and 70% of image depth, occluding either the region above or below it — 50 conditions per architecture. The two directions are complementary at a given boundary, so the fraction removed ranged from 30% to 70% in each direction.

Second, position was separated from amount. With the removed area fixed at 50% of image height, a contiguous band of that size was shifted across five vertical offsets spanning the image, so that the quantity of removed information was held constant while its location varied.

Third, the predicted probabilities were examined. For each model and condition we computed their standard deviation, interquartile range and number of distinct values (to three decimal places) across the 97 test images, with the expected calibration error [38] (15 equal-width confidence bins), the Brier score and the mean confidence on incorrect predictions — to distinguish chance-level discrimination arising from uncertain output from that arising from saturated output.

An AUC below 0.60 was used as an operational collapse criterion. Rates were also summarized at thresholds of 0.55, 0.65 and 0.70. Repeatability was checked for baseline and upper 50% occlusion within one computing environment. The original and follow-up inference implementations differed in batch size (64 versus 97 images), and the follow-up used a different computing environment; execution differences are reported in Supplementary Methods S12.

### 2.10 Label validation via blinded expert re-classification

To assess the anatomical validity of the acquisition-position labels used for training and evaluation, a label-validation sub-study was performed in which images were independently re-assigned to classes by blinded experts for comparison with the unchanged acquisition-position labels. Two examiners with anatomical expertise independently classified image subsamples as superior or inferior while blinded to the acquisition position and each other’s responses and rated their confidence on a 1–5 scale. The first examiner, who acquired the images, re-classified 20 unique images plus 5 duplicated images for intra-rater repeatability; because this examiner defined the original labels, that comparison is a label-grounding check rather than an independent test. A second anatomy expert, uninvolved in acquisition or labelling, independently classified 40 images, 20 of which overlapped with the first examiner’s set. Agreement was quantified as described in Section 2.11; analyses were repeated for the high-confidence subset (confidence ≥ 4).

### 2.11 Statistical analysis

Analyses were performed in Python. Training-seed means and standard deviations describe variation among five trained models on the fixed dataset. For the follow-up saved predictions at baseline and upper/lower 50% occlusion, 95% percentile confidence intervals were calculated by paired subject-cluster bootstrap [39] with 20,000 resamples (random seed 20261004). Eight test subjects were sampled with replacement, retaining all images of each selected subject with multiplicity. The same subject draws were used across architectures, training seeds and conditions; metrics were computed for each fixed trained seed and then averaged. Training seeds were not resampled. The AUC reduction was baseline AUC minus the mean of upper- and lower-occlusion AUC. These intervals quantify test-subject sampling uncertainty conditional on the saved models and predictions, not training-set uncertainty. Supplementary Table S14 reports these follow-up estimates separately from the original execution in Table 1. The original best-fold image-bootstrap intervals (1,000 resamples) and image-level Mann– Whitney, Wilcoxon, McNemar [40] and DeLong [41] analyses are retained only as exploratory summaries. They do not account for within-subject dependence and cannot establish population-level significance or equivalence. Holm adjustment was applied to the ten DeLong comparisons within each seed; intensity and ULR comparisons also used Holm and Benjamini–Hochberg adjustment. Adjustment does not correct clustering. Collapse counts across repeated conditions are reported descriptively, without Fisher testing. Expert agreement was summarized by percentage agreement and Cohen’s κ; repeat-inference rank agreement used Spearman’s ρ.

### 2.12 Use of artificial intelligence

The authors used Claude (Anthropic) for assistance with manuscript text, preliminary literature searches and summaries, analysis and figure-generation code, and reporting checklists, and Cursor (Anysphere) as an AI-assisted code editor. Codex (OpenAI) assisted with revision, consistency checks, subject-cluster bootstrap calculations from saved predictions, and submission-file preparation. These tools were used under the authors’ direction. The authors are responsible for reviewing and verifying the analyses and text and for the final content.

## 3. Results

### 3.1 Dataset characteristics and partitioning

Of 531 images acquired, 477 from 40 subjects met the inclusion criteria; 54 (10.2%) were excluded for file corruption, inadequate anatomical visualization or colour Doppler overlay. Classes were near-balanced (superior 240, 50.3%; inferior 237, 49.7%). All 40 subjects contributed to both classes within the same session, matching acquisition parameters across classes at subject level. Subject-independent partitioning yielded 380 training/validation images from 32 subjects and 97 held-out test images from 8 subjects (superior 49; inferior 48). The flow of participants and images is shown in Supplementary Figure S6.

### 3.2 Acquisition labels and expert agreement

Acquisition-position labels provided the reference for model training and testing. Expert re-classification assessed whether selected images could be assigned to the acquisition-defined classes from image appearance; it did not replace the acquisition labels or establish a pixel-level anatomical reference.

Labels were fixed by the position of the probe at acquisition and were therefore not derived from image appearance. Blinded expert re-classification nevertheless recovered them from the images alone (Supplementary Table S11). The acquiring examiner (an intra-rater, label-grounding check) agreed with the acquisition labels for 90.0% of 20 images (κ = 0.80) and for all high-confidence images (κ = 1.00). An independent expert, blinded to acquisition position, agreed for 85.0% of 40 images (κ = 0.70) and 96.7% of the 30 high-confidence images (κ = 0.93); on the 20 images both read, the two never disagreed with each other (κ = 1.00). Disagreement with the acquisition labels was almost entirely confined to low-confidence images, the one high-confidence exception being an image both experts independently assigned to the same opposite class. This disagreement may reflect image ambiguity or disagreement with the acquisition label; the reading study does not resolve its cause.

### 3.3 Classification performance

All five architectures were trained across five random seeds and evaluated on the same subject-independent held-out set (Table 1). Observed discrimination was similar across architectures: seed-mean AUC ranged from 0.958 ± 0.026 (ResNet-50) to 0.972 ± 0.008 (ConvNeXt-Tiny), and seed-mean F1 from 0.888 ± 0.029 (ResNet-50) to 0.911 ± 0.030 (DeiT-Tiny). The seed-to-seed standard deviations were of the same order as the differences between architectures. Figure 1 illustrates the separate original best-fold runs for three architectures; it does not depict the five-seed means.

**Figure 1.**
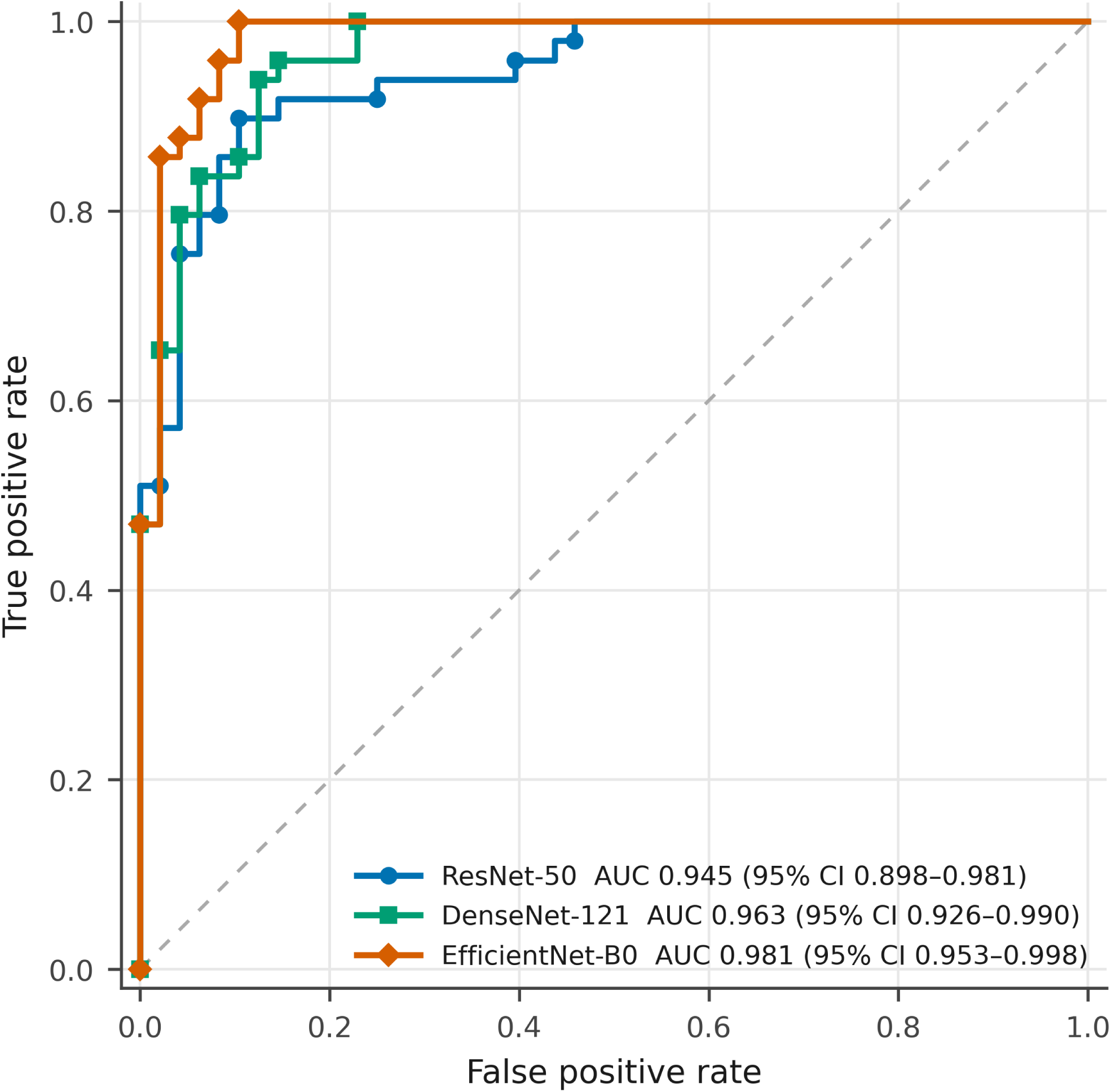
Receiver operating characteristic curves for the original single best-fold runs of ResNet-50, DenseNet-121 and EfficientNet-B0 on 97 test images from eight subjects. Legend intervals use 2,000 class-stratified image-bootstrap resamples (the archived ROC script), whereas Table 2 uses 1,000 resamples; small differences in interval limits are retained. These are exploratory 95% intervals and do not account for subject clustering. The curves are illustrative single-run results, not the five-seed means in Table 1. Subject-cluster intervals for the follow-up five-seed estimates are given in Supplementary Table S14. A receiver operating characteristic chart. The horizontal axis is false positive rate and the vertical axis true positive rate, both from 0 to 1. A dashed diagonal marks chance. Three stepped curves rise steeply toward the upper-left corner and lie close together. The legend gives each architecture with its area under the curve and exploratory image-bootstrap 95 per cent confidence interval for the original single run.

**Table 2.** Classification performance under occlusion ablation on the held-out test set (n = 97) for the single best-fold run of the three architectures carried through the detailed analyses. Ninety-five per cent confidence intervals were estimated by image-level bootstrap resampling (n = 1,000), without correction for within-subject clustering; these intervals are exploratory; class-wise accuracy is reported as n_correct/n_class. Upper-region occlusion replaces the upper half of the image; lower-region occlusion replaces the lower half. The occlusion boundary is a fixed geometric fraction of image height and is not anatomically delineated.

| **Model** | **Condition** | **Overall Acc (95% CI)** | **Inferior Acc** | **Superior Acc** | **F1 (95% CI)** | **AUC (95% CI)** |
|---|---|---|---|---|---|---|
| ResNet-50 | Baseline | 0.866 [0.794–0.928] | 0.812 (39/48) | 0.918 (45/49) | 0.874 [0.796–0.932] | 0.945 [0.895–0.981] |
| ResNet-50 | Upper-region occlusion | 0.639 [0.536–0.732] | 0.271 (13/48) | 1.000 (49/49) | 0.737 [0.645–0.816] | 0.874 [0.797–0.934] |
| ResNet-50 | Lower-region occlusion | 0.670 [0.577–0.763] | 0.417 (20/48) | 0.918 (45/49) | 0.738 [0.637–0.822] | 0.864 [0.773–0.938] |
| DenseNet-121 | Baseline | 0.907 [0.845–0.959] | 0.875 (42/48) | 0.939 (46/49) | 0.911 [0.844–0.962] | 0.963 [0.926–0.990] |
| DenseNet-121 | Upper-region occlusion | 0.660 [0.567–0.753] | 0.312 (15/48) | 1.000 (49/49) | 0.748 [0.661–0.824] | 0.897 [0.829–0.951] |
| DenseNet-121 | Lower-region occlusion | 0.804 [0.732–0.877] | 0.979 (47/48) | 0.633 (31/49) | 0.765 [0.657–0.857] | 0.888 [0.818–0.942] |
| EfficientNet-B0 | Baseline | 0.928 [0.876–0.969] | 0.917 (44/48) | 0.939 (46/49) | 0.929 [0.870–0.975] | 0.981 [0.951–0.998] |
| EfficientNet-B0 | Upper-region occlusion | 0.464 [0.371–0.567] | 0.083 (4/48) | 0.837 (41/49) | 0.612 [0.508–0.700] | 0.404 [0.317–0.498] |
| EfficientNet-B0 | Lower-region occlusion | 0.495 [0.392–0.588] | 0.208 (10/48) | 0.776 (38/49) | 0.608 [0.500–0.701] | 0.489 [0.402–0.571] |

EfficientNet-B7 was excluded after cross-validation and before test evaluation because it performed substantially worse and more variably (CV F1 = 0.630 ± 0.049), possibly reflecting overparameterization relative to dataset size (63.8 M parameters; 380 training images). Cross-validation F1 fell within ±0.08 of the test F1 for the three retained architectures with reported cross-validation results (Supplementary Table S5). Exploratory best-fold performance with image-bootstrap confidence intervals, precision, recall, confusion counts and inference latency (8.4–18.1 ms per image) is reported in Supplementary Table S10 and Supplementary Figure S2.

Exploratory image-level DeLong comparisons for all ten architecture pairs at five seeds are retained in Supplementary Table S13. One of 50 comparisons remained below 0.05 after within-seed Holm adjustment (ResNet-50 versus ConvNeXt-Tiny at seed 1; adjusted P = 0.025). Because images from the same subject are dependent, these P values are not used as confirmatory evidence of architecture differences.

The sign of the pairwise AUC difference reversed across seeds for all ten architecture pairs, and each architecture had the highest observed AUC at one seed. This describes variability in the ranking within this dataset; it does not establish equivalence. Subject-cluster intervals for the follow-up baseline predictions are reported in Supplementary Table S14.

### 3.4 Performance under synthetic occlusion

Under the original 50% occlusion evaluation, four architectures retained AUCs of 0.851–0.917, with mean reductions of 0.063–0.109, whereas EfficientNet-B0 reached 0.549 and 0.543 under upper and lower occlusion (mean reduction 0.417 ± 0.044; Table 1). Both occluded AUCs were below 0.60 in each of its five seeds. Detailed best-fold results are reported in Table 2 as exploratory single-run summaries. In the follow-up execution, EfficientNet-B0 had mean AUCs of 0.510 and 0.522; its mean reduction was 0.447 (subject-cluster 95% CI 0.396–0.495), compared with 0.063–0.109 for the other architectures (Supplementary Table S14). The numerical discrepancy and execution differences are described in Supplementary Methods S12.

Class-wise responses were largely shared. Across seeds, all four architectures that retained discrimination defaulted toward the superior class when the upper region was occluded (superior accuracy 0.931–1.000; Figure 2B). Under lower-region occlusion no consistent default emerged, and the inverted default reported for DenseNet-121 in a single best-fold run (inferior accuracy 0.979) lay at the extreme of the seed distribution (0.658 ± 0.269), so we do not interpret it as a property of dense connectivity. EfficientNet-B0 lost accuracy in both classes under both conditions and showed no coherent default at all.

**Figure 2.**
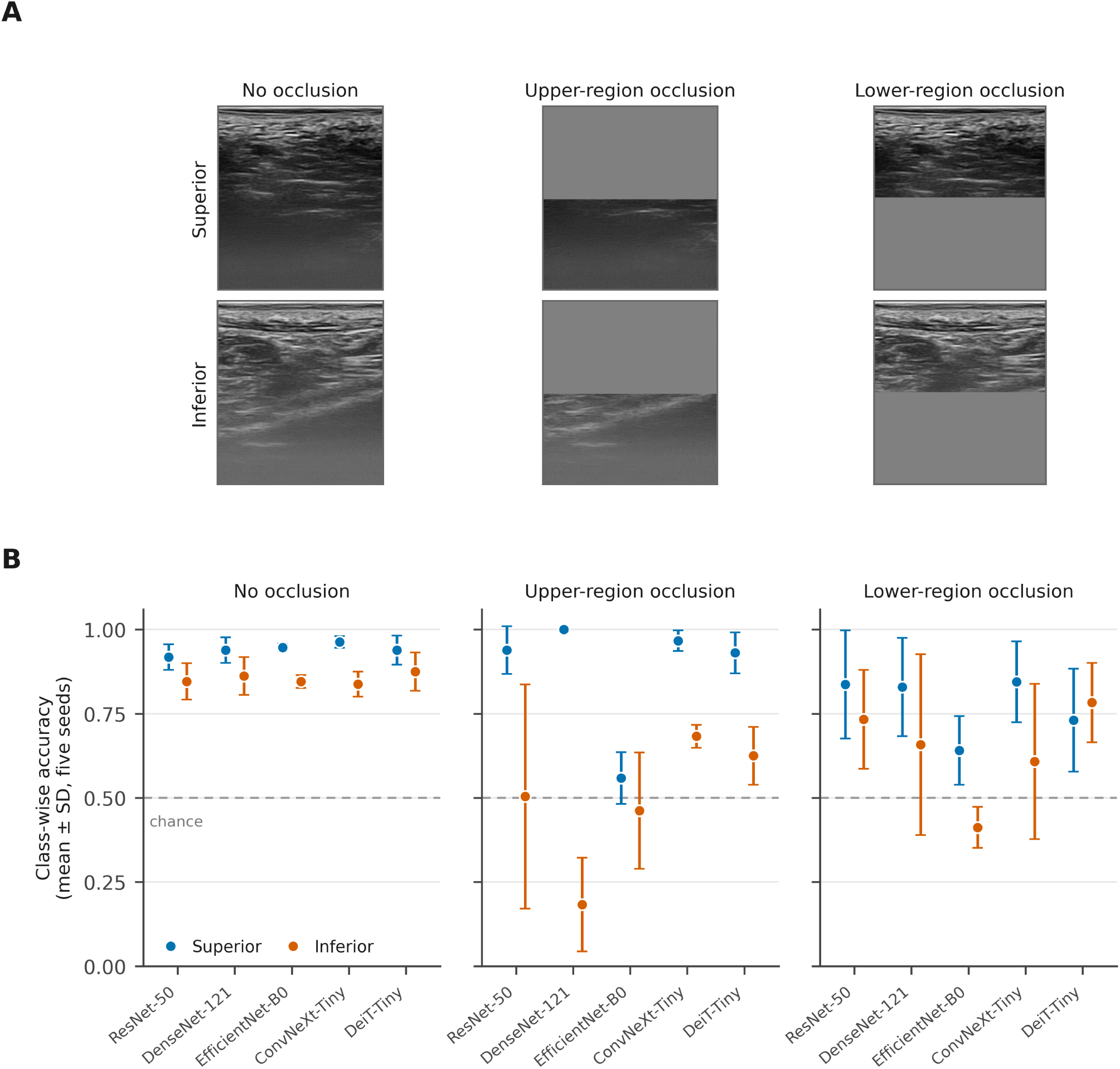
Occlusion ablation. (A) Representative occluded image pairs for the superior and inferior classes: baseline, upper-region occlusion and lower-region occlusion, each replaced with the training-set mean intensity. (B) Class-wise accuracy under each condition, shown as the mean ± SD across five random seeds for all five architectures. Values derive from the follow-up saved predictions. Dotted line indicates chance (0.50). Four architectures default toward the superior class under upper-region occlusion; EfficientNet-B0 degrades in both classes under both conditions. Panel A shows six greyscale intraoral ultrasound images in two rows and three columns. The top row is a superior-class image and the bottom row an inferior-class image; the left column is unaltered, the middle column has its upper half replaced by flat grey, and the right column has its lower half replaced by flat grey. Panel B shows three dot-and-error-bar charts of class-wise accuracy for five architectures, one chart per occlusion condition, with a dotted line marking chance at 0.50.

Because the 50% boundary is arbitrary, each seed model was re-evaluated with the boundary at 30%, 40%, 50%, 60% and 70% of image depth, occluding the region above or below it (50 conditions per architecture; Figure 3). EfficientNet-B0 fell below an AUC of 0.60 in 49 of 50 conditions (mean 0.494; minimum 0.357), whereas ResNet-50 and ConvNeXt-Tiny did so in none, DenseNet-121 in 1 of 50 and DeiT-Tiny in 2 of 50; these three exceptions all occurred at the largest removal fraction. The four architectures showed an overall decline in discrimination as the fraction removed increased and retained an AUC of 0.917–0.956 when 30% was removed, whereas EfficientNet-B0 was already at chance at that fraction (0.520 and 0.498 for upper- and lower-region occlusion). These repeated-condition counts are descriptive; no test treating the conditions as independent observations was used. The result was also robust to the collapse criterion: across thresholds from 0.55 to 0.70, EfficientNet-B0’s collapse rate ranged from 0.82 to 1.00 and no other architecture exceeded 0.12.

**Figure 3.**
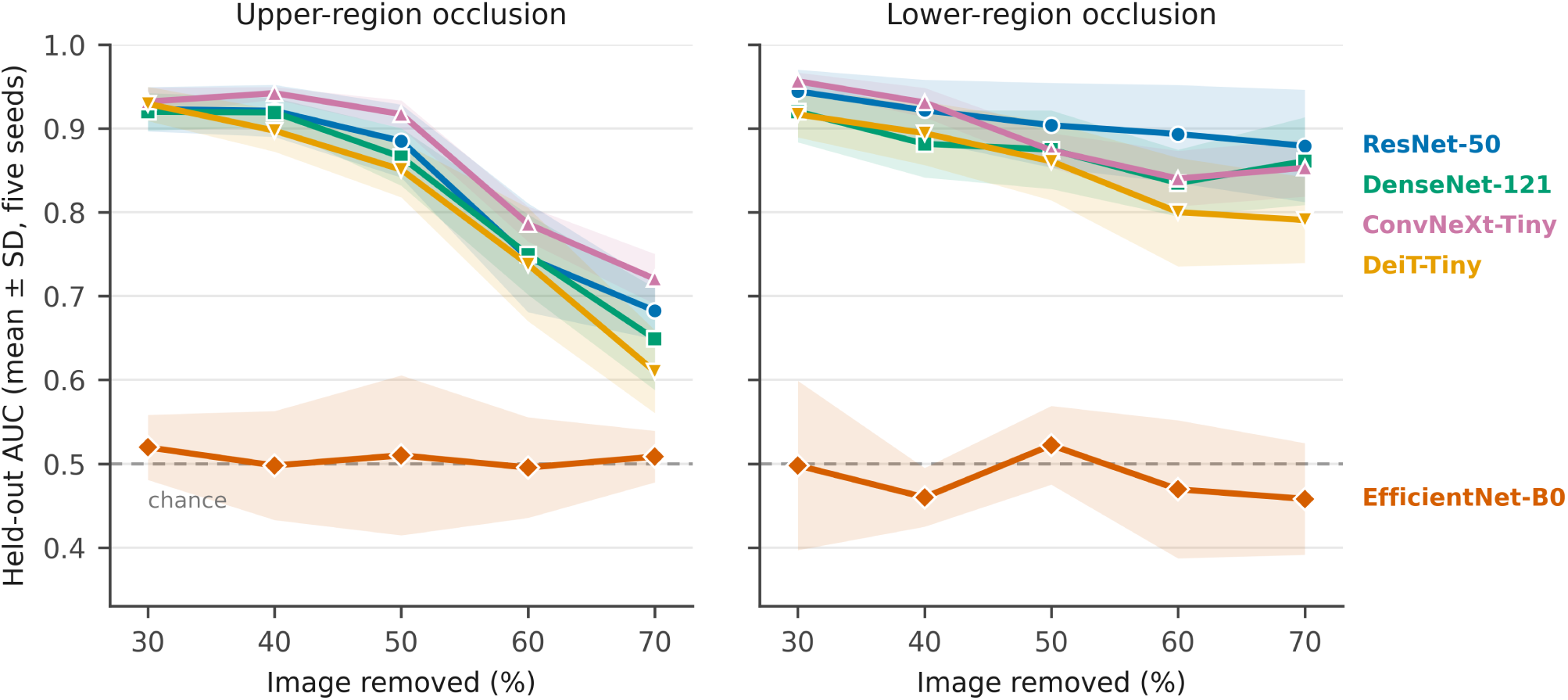
Occlusion sensitivity across architectures and boundary positions. Held-out AUC (mean ± SD across five random seeds) when the upper (left panel) or lower (right panel) region of each test image was replaced with the training-set mean, plotted against the fraction of the image removed. Boundaries were placed at 30–70% of image depth. Dashed line denotes chance (AUC 0.50). Four architectures show an overall decline as the removed fraction increases; EfficientNet-B0 has mean AUC near 0.50 across the tested fractions. Shaded bands describe seed variation, not subject-sampling uncertainty. Two line charts side by side. The horizontal axis is the fraction of the image removed, from 30 to 70 per cent; the vertical axis is area under the curve, with chance at 0.50. Each chart plots five architectures as separate lines with distinct markers. In both charts four lines slope gently downward and stay well above chance, while a fifth line remains near chance across the tested range.

In the area-matched control, a band covering 50% of image height was shifted across five vertical offsets. EfficientNet-B0 had a mean AUC of 0.503 (minimum 0.404), while mean AUCs for the other architectures were 0.811–0.869. The observed sensitivity therefore extended across the tested mask positions at a fixed area. This does not exclude sensitivity to the mask itself or establish independence from mask size outside the tested range.

Among the architectures that retained discrimination, occluding the upper region degraded performance more than occluding the lower region at matched removal fractions (at 70% removal, AUC 0.610–0.720 versus 0.791– 0.879). Because the boundary is geometric rather than anatomically delineated, we report this asymmetry descriptively only.

### 3.5 Discrimination and confidence under occlusion

The loss of discrimination in EfficientNet-B0 was accompanied by output saturation rather than a general shift toward uncertain predictions. Under 50% occlusion, its predicted probabilities took only 8–15 distinct values (to three decimal places) across the 97 test images, compared with approximately 91 when unoccluded, and their interquartile range approached the maximum possible (0.998–1.000). The spread of its outputs increased relative to baseline (SD ratio 1.34), whereas every other architecture showed the opposite (0.47–0.83) (Supplementary Table S12; Supplementary Figure S5). These near-binary predictions had markedly reduced input-specific variation. AUC may be sensitive to small differences among saturated probabilities; the contribution of numerical effects was not isolated.

This was accompanied by extreme confidence. Calibration and confidence were quantified for every seed model at baseline and under occlusion (Table 3). On intact images, expected calibration error ranged from 0.061 to 0.094 across the five architectures, and mean confidence on incorrect predictions ranged from 0.644 to 0.783; EfficientNet-B0 was the least confident of the five when wrong (0.644). Occlusion reversed this ordering: its accuracy fell to 0.520, its confidence on incorrect predictions rose to 0.989 (other architectures 0.714–0.776) and its expected calibration error to 0.478 (0.095–0.199), and 92.9% of its predicted probabilities were below 0.01 or above 0.99. The lowest baseline confidence on errors did not predict the high confidence observed on errors under synthetic occlusion.

**Table 3.** Calibration and confidence at baseline and under occlusion, averaged across five random seeds in the follow-up execution (Supplementary Methods S12). Expected calibration error was computed with 15 equal-width bins. Occluded values are averaged over upper- and lower-region occlusion at a boundary of 50% of image depth. Extreme probabilities are the proportion of predicted probabilities below 0.01 or above 0.99.

| **Architecture** | **Baseline ECE** | **Baseline confidence when incorrect** | **Occluded accuracy** | **Occluded ECE** | **Occluded confidence when incorrect** | **Occluded extreme probabilities** |
|---|---|---|---|---|---|---|
| ResNet-50 | 0.072 | 0.783 | 0.755 | 0.153 | 0.776 | 25.7% |
| DenseNet-121 | 0.075 | 0.739 | 0.670 | 0.199 | 0.756 | 13.0% |
| EfficientNet-B0 | 0.094 | 0.644 | 0.520 | 0.478 | 0.989 | 92.9% |
| ConvNeXt-Tiny | 0.061 | 0.757 | 0.777 | 0.117 | 0.758 | 5.1% |
| DeiT-Tiny | 0.072 | 0.767 | 0.768 | 0.095 | 0.714 | 0.6% |

### 3.6 Grad-CAM attribution: reproducibility and faithfulness

Grad-CAM spatial metrics — mean activation intensity, normalized vertical centroid and the upper-to-lower activation ratio — were recomputed across the five random seeds and across three alternative class-activation methods (Grad-CAM++, Score-CAM, Layer-CAM). Neither the magnitude nor, for several architecture–metric pairs, the direction of the between-class difference was preserved; EfficientNet-B0’s centroid pattern reversed under Score-CAM (Supplementary Tables S7 and S8). Single-run values are therefore reported descriptively in Table 4 and Figures 4 and 5 and are not interpreted as evidence of anatomical alignment.

**Figure 4.**
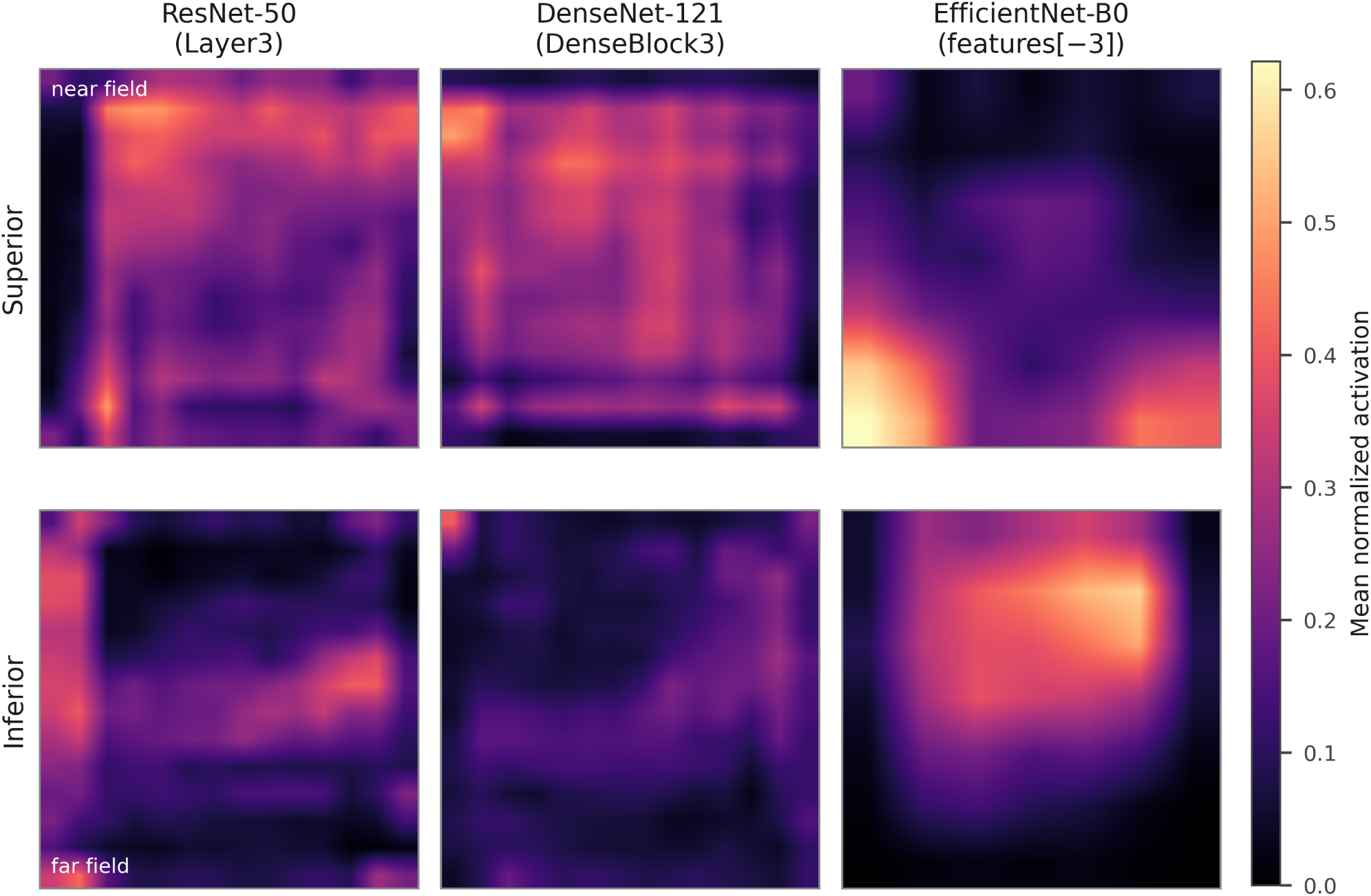
Average Grad-CAM activation maps aggregated by class (superior, top row; inferior, bottom row) for each architecture carried through the detailed analysis, with the target layer given above each column. Each map is the pixel-wise mean, over all test images of a class, of the per-image Grad-CAM heatmaps, each min–max normalized to [0, 1]; the vertical axis corresponds to imaging depth (top = near field, bottom = far field). A perceptually uniform colour scale is used so that equal steps in activation appear as equal steps in colour, and the same scale applies to all three architectures. These maps are from a single run and are shown descriptively. Six square activation maps in two rows and three columns, one column per architecture and one row per class, on a dark-to-light colour scale with a shared scale bar at the right. The vertical axis of each map corresponds to imaging depth, labelled near field at the top and far field at the bottom. Bright regions mark higher mean activation; their position differs between the two rows for the third column and much less so for the first two.

**Figure 5.**
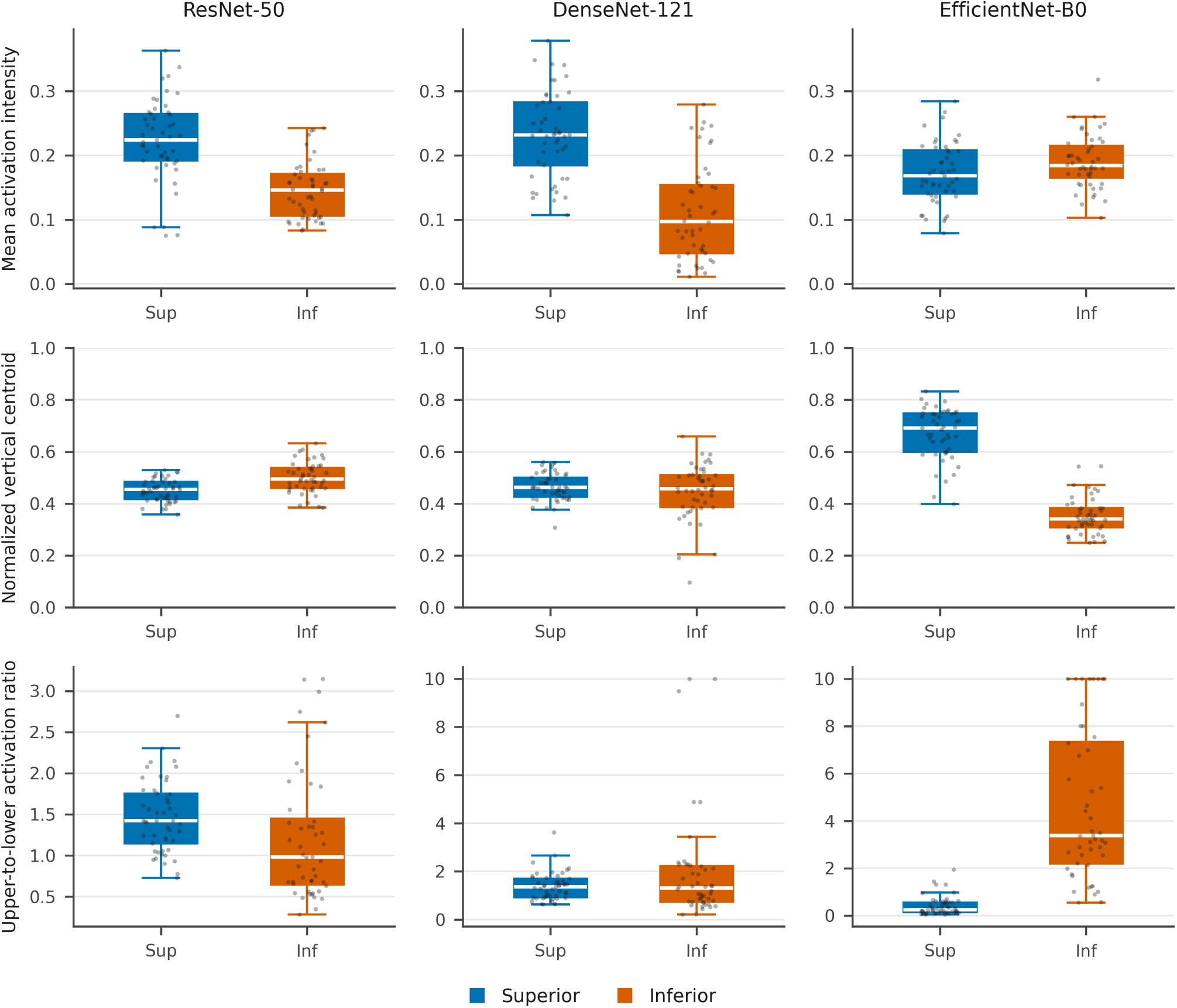
Class-wise distributions of three Grad-CAM spatial metrics — mean activation intensity (top row), normalized vertical centroid (middle row) and upper-to-lower activation ratio (bottom row) — for each architecture carried through the detailed analysis. Boxes show the median and interquartile range with individual images overlaid; intensity shares a common scale across architectures, the activation ratio does not. Exploratory image-level between-class comparisons, without subject-cluster adjustment, used the two-sided Mann–Whitney U test (Table 4). These single-run patterns varied across seeds or class-activation methods (Supplementary Table S7). A three-by-three grid of box plots with individual data points overlaid. Rows are three attribution metrics and columns are three architectures. Within each panel the two boxes are the superior and inferior classes in two colours. The top row shares a common vertical scale across columns; the bottom row does not.

**Table 4.**
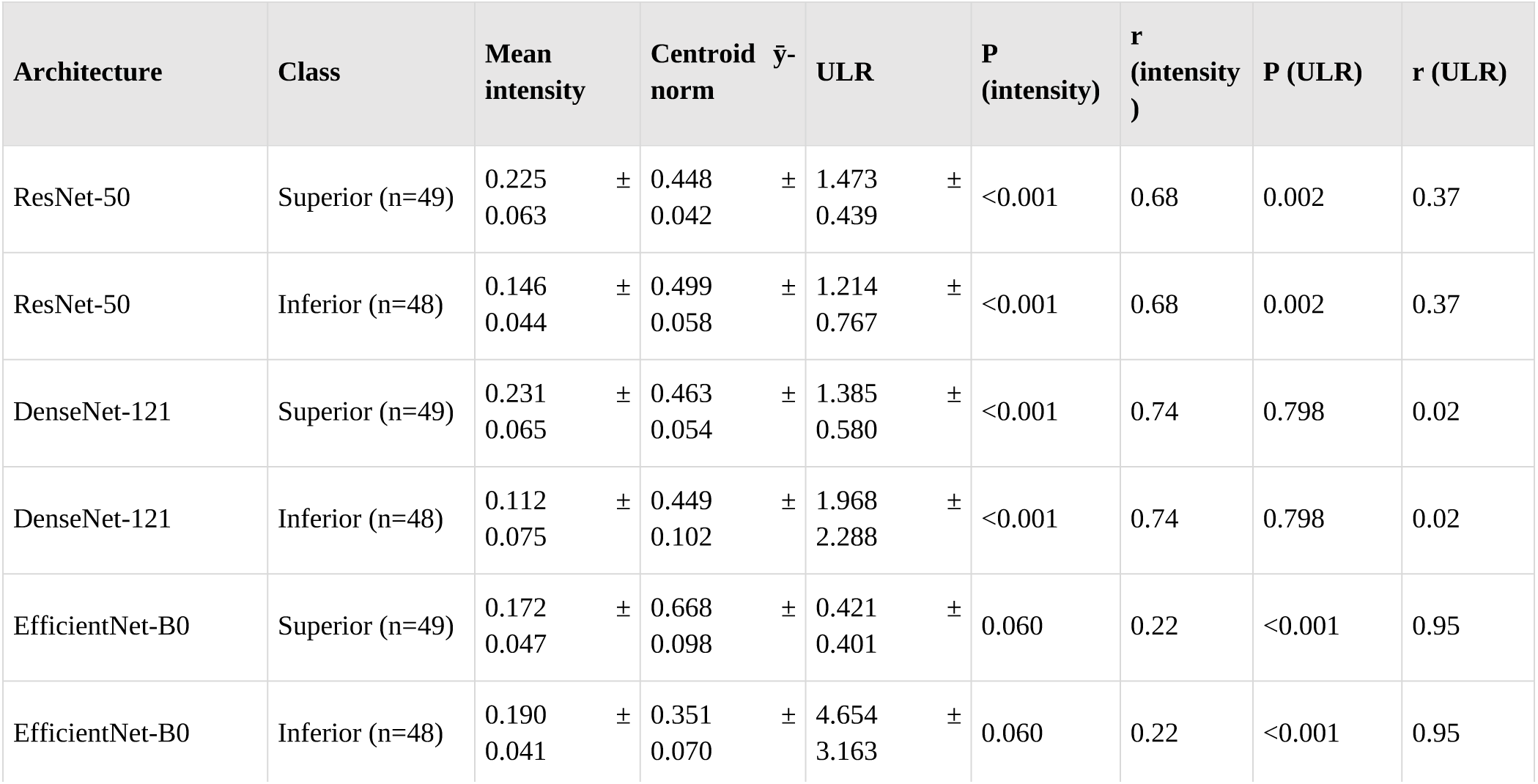
Quantitative Grad-CAM spatial metrics by class, single run. Mean activation intensity, normalized vertical centroid (0 = image top, 1 = image bottom) and upper-to-lower activation ratio (ULR), each mean ± SD. ULR was clipped at 10.0 to suppress numerical instability from near-zero lower activation. These single-run patterns varied across seeds or class-activation methods (Supplementary Table S7) and are reported descriptively. Image-level P values do not account for subject clustering and are exploratory.

| **Architecture** | **Class** | **Mean intensity** | **Centroid ȳ-norm** | **ULR** | **P (intensity)** | **r (intensity)** | **P (ULR)** | **r (ULR)** |
|---|---|---|---|---|---|---|---|---|
| ResNet-50 | Superior (n=49) | 0.225 ± 0.063 | 0.448 ± 0.042 | 1.473 ± 0.439 | <0.001 | 0.68 | 0.002 | 0.37 |
| ResNet-50 | Inferior (n=48) | 0.146 ± 0.044 | 0.499 ± 0.058 | 1.214 ± 0.767 | <0.001 | 0.68 | 0.002 | 0.37 |
| DenseNet-121 | Superior (n=49) | 0.231 ± 0.065 | 0.463 ± 0.054 | 1.385 ± 0.580 | <0.001 | 0.74 | 0.798 | 0.02 |
| DenseNet-121 | Inferior (n=48) | 0.112 ± 0.075 | 0.449 ± 0.102 | 1.968 ± 2.288 | <0.001 | 0.74 | 0.798 | 0.02 |
| EfficientNet-B0 | Superior (n=49) | 0.172 ± 0.047 | 0.668 ± 0.098 | 0.421 ± 0.401 | 0.060 | 0.22 | <0.001 | 0.95 |
| EfficientNet-B0 | Inferior (n=48) | 0.190 ± 0.041 | 0.351 ± 0.070 | 4.654 ± 3.163 | 0.060 | 0.22 | <0.001 | 0.95 |

Single-run class differences varied by architecture and metric (Table 4). Across all five seeds, the majority direction occurred in three of five runs for ResNet-50, DenseNet-121, EfficientNet-B0 and DeiT-Tiny, and four of five for ConvNeXt-Tiny, for each of the three spatial metrics (Supplementary Table S7). These counts use all seeds, irrespective of image-level P values. They describe limited directional consistency in this sample rather than a validated threshold for attribution stability.

In the detailed single-run insertion analysis, Grad-CAM and random insertion AUCs were 0.791 versus 0.583 for ResNet-50, 0.731 versus 0.546 for DenseNet-121, and 0.546 versus 0.532 for EfficientNet-B0 (Table 5). The original image-level Wilcoxon comparison for EfficientNet-B0 did not detect a difference (P = 0.22); this does not establish equivalence to random attribution or absence of class-relevant information. These comparisons do not account for subject clustering. ConvNeXt-Tiny and DeiT-Tiny had positive observed insertion gain at all five seeds; deletion AUC and Average Drop were not computed for them.

**Table 5.** Insertion and deletion evaluation of Grad-CAM attributions. Insertion and deletion area under the curve for Grad-CAM attributions versus random masking, and Average Drop. Exploratory final column, unadjusted for subject clustering: two-sided Wilcoxon signed-rank comparison of per-image insertion gain (attribution versus random).

| **Model** | **Insertion AUC (Grad-CAM)** | **Insertion AUC (random)** | **Deletion AUC** | **Average Drop (%)** | **Grad-CAM vs. random (Wilcoxon p)** |
|---|---|---|---|---|---|
| ResNet-50 | 0.791 | 0.583 | 0.460 | 33.9 | 1.0 × 10<sup>−15</sup> |
| DenseNet-121 | 0.731 | 0.546 | 0.479 | 37.4 | 4.3 × 10<sup>−17</sup> |
| EfficientNet-B0 | 0.546 | 0.532 | 0.511 | 33.9 | 0.22 |

### 3.7 Misclassified cases

Representative correctly classified and misclassified cases are shown in Figure 6 and Supplementary Figure S3. Selected errors showed indistinct fat-pad–muscle boundaries or a weak or atypically positioned dermis echo. These qualitative observations suggest possible sources of difficulty but do not establish that anatomical ambiguity explains all errors or that error mechanisms are shared across architectures.

**Figure 6.**
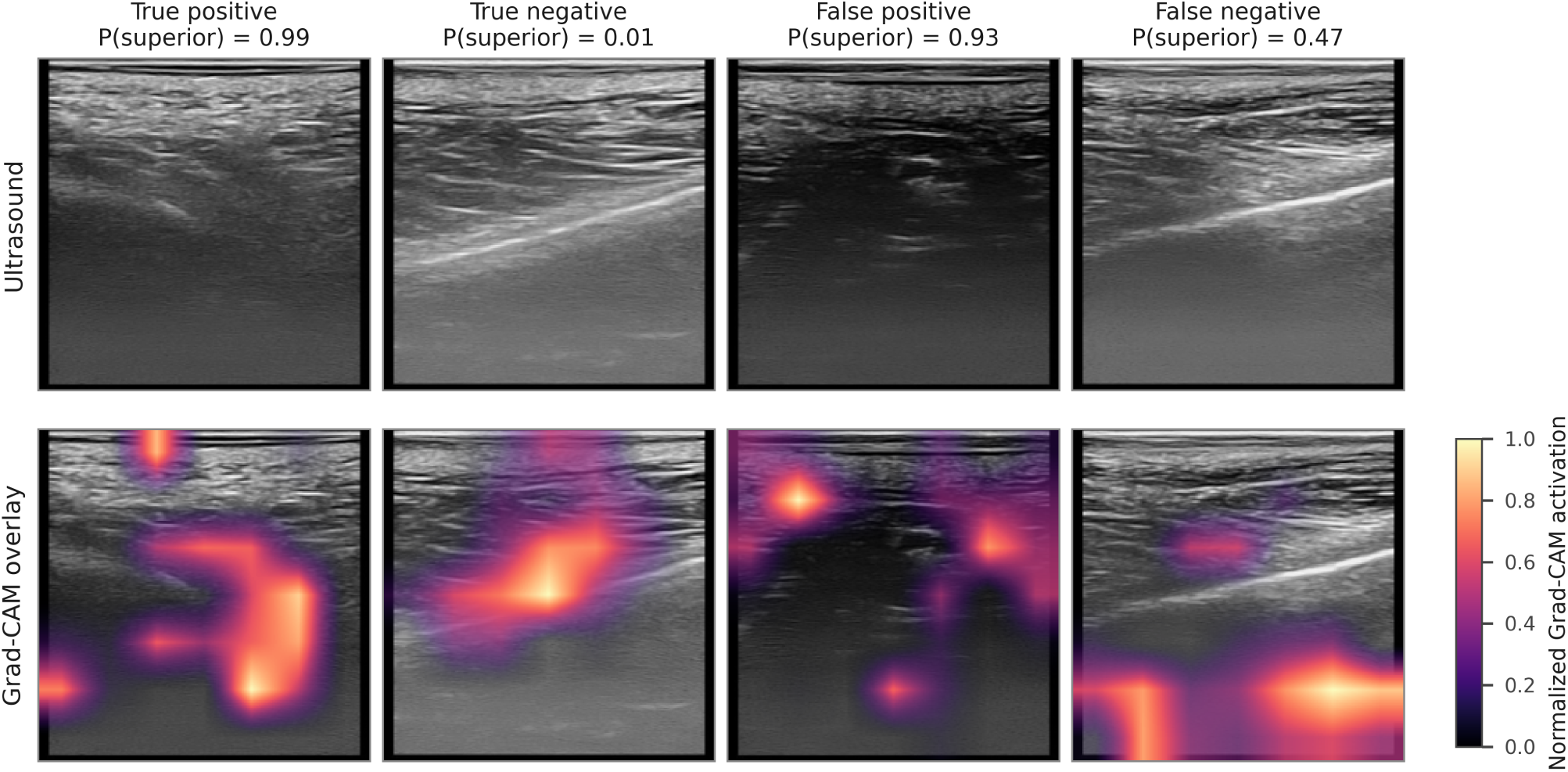
Representative correctly classified (true positive and true negative) and misclassified (false positive and false negative) cases from EfficientNet-B0 on the held-out test set, selected by the highest confidence within each prediction category, with the predicted probability of the superior class above each column. Top row: original ultrasound images; bottom row: Grad-CAM overlays on the same images, on a perceptually uniform colour scale with low activation left transparent. The overlays illustrate the appearance of these cases and, for the reasons given in Section 3.6, are not offered as evidence of what the model used. Eight images in two rows and four columns. The top row shows four greyscale intraoral ultrasound images labelled true positive, true negative, false positive and false negative, each with the model’s predicted probability above it. The bottom row repeats the same four images with a colour attribution overlay in which low activation is transparent, alongside a scale bar.

## 4. Discussion

### 4.1 Principal findings

This study examined how models with similar baseline discrimination differed when evaluated through perturbation responses, confidence and spatial attribution. Acquisition-position classification provided a tractable test case in intraoral ultrasonography without requiring a diagnostic claim. Expert agreement supported the interpretability of selected acquisition-defined labels, but neither the labels nor expert reading supplied a complete reference for the image features a model should use.

The central finding was a mismatch between baseline performance and behaviour under altered inputs. Five architectures achieved similar observed baseline discrimination, yet EfficientNet-B0 fell below AUC 0.60 in 49 of 50 boundary-sweep conditions and remained sensitive in the area-matched control. Its loss of discrimination was accompanied by highly confident errors rather than reduced confidence. Spatial attribution patterns also varied across seeds and methods, limiting their ability to resolve the interpretation of model behaviour. The contribution is therefore the joint characterization of these discrepancies in intraoral ultrasonography, rather than a general assertion that accuracy alone is insufficient. The experiments do not identify their anatomical or architectural mechanism.

### 4.2 Similar baseline discrimination and different perturbation responses

Seed-mean AUC ranged from 0.958 to 0.972 and F1 from 0.888 to 0.911. Pairwise rankings reversed across seeds, and the eight-subject test cohort leaves substantial uncertainty. Non-significant exploratory comparisons cannot demonstrate equivalence. Architecture choice based on a single run would not capture the observed variability or the differences under synthetic perturbation.

EfficientNet-B7 performed poorly in cross-validation before test evaluation. Its larger capacity is one possible explanation, but overparameterization was not tested directly. Differences between cross-validation and test rankings may also reflect the small subject counts in the folds (Section 4.7).

EfficientNet-B0 had a mid-range baseline AUC but the largest observed loss under the tested masks. Boundary sweeps showed that the response was not confined to the primary 50% boundary, and the area-matched control showed that it persisted across mask positions when the removed area was fixed. The resulting distinction between evaluated pipelines was largely absent from the baseline performance comparison. This supports considering perturbation response as an additional dimension of model comparison, while retaining the uncertainty from the small test cohort and the differences in training settings. It does not establish an intrinsic robustness ranking of architecture families.

### 4.3 Occlusion sensitivity and possible explanations

Attribution maps do not by themselves establish causal use of the highlighted regions [42]. Constant-intensity occlusion changes both image content and the input distribution. The retained AUC of four architectures under 50% occlusion shows discrimination under those masked inputs; the larger loss for EfficientNet-B0 shows sensitivity to the intervention. Boundary and area controls extend this observation across specified locations and removal fractions, but do not separate dependence on distributed anatomical information from susceptibility to the artificial mask.

Potential explanations include different dependence on spatially distributed image information and different susceptibility to the artificial mask. Residual propagation [24], depthwise-separable convolutions and squeeze- and-excitation weighting [26] provide architectural context for generating hypotheses, but this experiment did not isolate their effects. Differences in tuning, optimization and execution also prevent attributing the observed response to a particular design component. The most defensible interpretation is a difference among the evaluated model pipelines under the specified perturbations.

Class-wise responses were largely shared (Section 3.4). The inverted default reported for DenseNet-121 in a single best-fold run lay at the extreme of the seed distribution, so we do not interpret it as a property of dense connectivity — an instance of the same single-run instability this study documents for attribution.

### 4.4 Loss of discrimination without reduced confidence

The confidence response gives the occlusion result a separate implication. EfficientNet-B0 had the lowest mean confidence on incorrect predictions at baseline, yet under 50% occlusion its accuracy fell to 0.520 while mean confidence on incorrect predictions rose to 0.989. It assigned 92.9% of probabilities below 0.01 or above 0.99. Thus, favourable confidence behaviour on intact images did not carry over to these altered inputs. A review strategy that flags only low-confidence predictions could miss high-confidence errors of this type, although no clinical triage rule or threshold was evaluated here.

Expected calibration error increased from 0.094 on intact images to 0.478 under occlusion. Evaluating confidence together with discrimination under the same perturbation therefore revealed information that either baseline calibration or perturbed AUC alone would miss. Probe angulation, acoustic coupling and field of view vary in clinical imaging, but constant masks do not reproduce these changes. The observed discrimination–confidence mismatch motivates testing whether confidence remains informative under realistic acquisition variation; it does not demonstrate that the same failure occurs clinically.

### 4.5 Attribution reproducibility as a constraint on interpretation

Attribution adds a third dimension to this evaluation, but did not provide a stable anatomical account of the observed model differences. The direction and magnitude of Grad-CAM metrics varied across seeds and methods. For EfficientNet-B0, the small observed insertion advantage over random masking was not detected as significant in the original image-level test. That result is inconclusive about equivalence and does not prove that the maps contain no relevant information. Instead, seed and method variation, together with the limited insertion evidence, constrain what can be inferred from a plausible single-run map. Reproducibility and faithfulness address different aspects of attribution and should not be inferred from visual plausibility alone.

Benchmarking studies elsewhere in medical imaging have reported that saliency maps localize abnormalities less accurately than expert readers [43] and can fail model-randomization and repeatability checks [44].

### 4.6 Misclassification patterns

Selected misclassified examples showed indistinct tissue boundaries or a weak dermis echo. Such appearances provide plausible explanations for reader and model difficulty, but a systematic causal link between ambiguity and model error was not established. These examples are descriptive and do not validate an automated referral rule.

### 4.7 Methodological considerations

Hyperparameter optimization used a single validation fold rather than nested cross-validation, admitting fold-specific selection bias [32]. ConvNeXt-Tiny and DeiT-Tiny also used different optimizer search spaces, and DeiT-Tiny used a lower learning-rate range. Architecture comparisons therefore reflect the evaluated training pipelines as well as model design; they do not isolate architecture as the sole cause of performance differences.

The primary analyses average five training seeds on the saved Fold 2 partition (Table 1). For ResNet-50, DenseNet-121 and EfficientNet-B0, the detailed best-fold run is reported separately (Table 2; Supplementary Tables S5 and S10); Fold 2 was selected after tuning on Fold 1. For ConvNeXt-Tiny and DeiT-Tiny, both tuning and early stopping used Fold 2. The held-out test subjects were separate from these training/validation partitions, but seed replication does not remove single-fold model-selection bias.

All images were acquired by one operator. This reduces between-operator variation within the dataset but limits generalizability and may allow operator-specific features to influence classification. The measured acquisition-parameter comparisons did not establish the absence of confounding.

Finally, the upper-to-lower activation ratio was introduced to separate spatial from amplitude-related differences in attention. Because it was not reproducible here, we offer it as a diagnostic template whose outputs must themselves be checked for stability before anatomical interpretation.

### 4.8 Implications for future intraoral ultrasound models

The relevance to dentomaxillofacial imaging lies in comparing models and interpreting their outputs before considering diagnostic or procedural use. Similar discrimination on intact images may coexist with different sensitivity to altered inputs, and a model may remain highly confident as discrimination deteriorates. An attribution overlay provides further evidence only to the extent that its patterns are reproducible and supported by the evaluation used. The present subregion-classification task demonstrates these evaluation issues within a limited intraoral ultrasound setting.

A useful evaluation would report discrimination and confidence together on both intact and clinically motivated altered inputs, then examine attribution reproducibility across training runs and methods and its performance against appropriate controls. Such reporting could distinguish loss of discrimination accompanied by uncertainty from loss accompanied by confident errors, while making the limits of attribution-based interpretation explicit. Stored checkpoints allow some of these checks without retraining, although realistic acquisition perturbations and independent validation require additional work. These checks complement clinical validation and do not establish safety by themselves.

The present findings apply to the sampled buccal subregion task, evaluated training pipelines and specified masks. Their value is in exposing differences between dimensions of model behaviour that would be obscured by a comparison based only on baseline discrimination.

### 4.9 Limitations and future directions

The dataset comprises 477 images from 40 subjects at one institution, with only eight subjects in the held-out test set. The subject-cluster bootstrap addresses within-subject dependence for the saved follow-up predictions but remains imprecise with so few clusters and conditions on the observed training data and fixed models. Seed replication does not add independent participants. Earlier image-level intervals and tests are exploratory and may be anticonservative. Generalization beyond this cohort requires evaluation with additional centres and operators.

Expert re-classification supported agreement with acquisition-position labels in selected images, not pixel-level correspondence between attribution and anatomy. Anatomical regions were not delineated. Direct comparison against expert regions, alternative perturbation methods and model-randomization controls could provide additional evidence in future work.

Constant-intensity masks are artificial inputs. Their effects may reflect loss of distributed information, distribution shift or both. Geometric boundaries do not isolate individual tissue layers. Differences between original and follow-up EfficientNet-B0 occluded AUCs also remain unexplained; changing batch size and environment are possible contributors, not demonstrated causes. Repeatability of baseline and upper-mask predictions within one environment does not resolve this between-execution discrepancy.

Finally, this study addressed binary subregion classification, not vessel detection, flap viability assessment or surgical risk stratification; its results are not evidence for surgical decision support. The possible explanations in Section 4.3 remain hypotheses. The confidence-based review implications in Section 4.4 were not evaluated as a clinical workflow.

Future work could evaluate clinically motivated acquisition changes and extend perturbation assessment to continuous anatomical measurements. A lack of output change after occlusion would need to be interpreted alongside redundant information and distribution shift rather than treated as proof that the region was unused.

## 5. Conclusions

In intraoral ultrasound subregion classification, similar observed baseline discrimination concealed different responses to synthetic occlusion. Across five training seeds, EfficientNet-B0 fell below AUC 0.60 in 49 of 50 boundary-sweep conditions, while its errors under 50% occlusion became highly confident. Spatial attributions were not consistently reproduced across seeds or class-activation methods, limiting anatomical interpretation from a single map. Joint reporting of perturbation sensitivity, confidence under perturbation and attribution reproducibility therefore characterized model behaviour beyond baseline discrimination. These findings do not isolate architecture as the cause or predict clinical performance; confirmation requires additional participants and realistic acquisition conditions.

## Data Availability

The ultrasound images are not publicly available because the institutional review board approval under which they were acquired does not extend to public release. De-identified derived data supporting the findings are available from the corresponding author on reasonable request. Analysis code for the occlusion, sensitivity and attribution experiments is available from the corresponding author on request and will be deposited in a public repository with a persistent identifier.

## Supplementary material

Supplementary Methods S1–S13; Supplementary Tables S1–S14; Supplementary Figures S1–S6. Table S13 retains exploratory image-level DeLong comparisons; Table S14 reports follow-up subject-cluster uncertainty estimates.

## Data and code availability

Analysis code for the occlusion, sensitivity and attribution experiments is available from the corresponding author on request and will be deposited in a public repository with a persistent identifier. De-identified derived data supporting the findings are available from the corresponding author on reasonable request. The ultrasound images are not publicly available because the institutional review board approval under which they were acquired does not extend to public release.

## Declarations

### Ethics approval and consent to participate

This study was approved by the Institutional Review Board of Yonsei University Dental Hospital (approval no. 2-2023-0050; approved on 19 October 2023) and was conducted in accordance with the Declaration of Helsinki. All participants provided written informed consent.

### Funding

This work was supported by the National Research Foundation of Korea (RS-2024-00406281). The funder had no role in the design of the study; in the collection, analysis or interpretation of the data; in the writing of the manuscript; or in the decision to publish the results.

### Competing interests

The authors declare no conflict of interest.

### Author contributions (CRediT)

Goun Jung: Conceptualization, Methodology, Software, Formal analysis, Investigation, Data curation, Writing — original draft, Visualization. Yu-Ran Heo: Methodology, Investigation, Resources, Data curation, Writing — review and editing. Kyung-Seok Hu: Validation, Writing — review and editing. Hee-Jin Kim: Conceptualization, Resources, Supervision, Project administration, Funding acquisition, Writing — review and editing.

### Use of artificial intelligence

The use of AI tools is described in Section 2.12.

## Notes

### Competing Interest Statement

The authors have declared no competing interest.

### Author Declarations

The study was approved by the Institutional Review Board of Yonsei University Dental Hospital (approval no. 2-2023-0050; approved on 19 October 2023). All participants provided written informed consent.

