## Supplementary Material for "Similar discrimination, different behaviour under synthetic occlusion: confidence and attribution reproducibility of deep learning models for intraoral ultrasonography"

### Supplementary Methods

#### S1. Ultrasound Imaging Protocol

##### S1.1 Equipment

All intraoral ultrasound examinations were performed using the ECUBE PLATINUM ultrasound system (Alpinion Medical Systems, Seoul, Republic of Korea) equipped with an IO3-12 high-frequency intraoral linear transducer. The operating frequency was set to 11.5 MHz and acoustic power output was maintained at 100% for all acquisitions, with no variation across subjects or scanning positions.

##### S1.2 Scanning Protocol

All examinations were performed by a single trained operator to minimize inter-operator variability in probe placement and contact pressure. The transducer was oriented medial-superior and positioned in contact with the buccal mucosa without compression. For each subject, images were acquired at both Superior and Inferior scanning positions during the same examination session. Superior buccal mucosa images were acquired with the probe positioned at the upper buccal subregion overlying the zygomaticus major and levator anguli oris musculature. Inferior buccal mucosa images were acquired at the lower subregion overlying the buccinator muscle belly.

##### S1.3 Adjustable Parameters and Acquisition Rationale

Imaging preset, dynamic range, gain, and imaging depth were selected by the operator on a per-subject basis to optimize visualization of the target tissue layers, in accordance with standard clinical practice for intraoral ultrasonography. Fixing these parameters uniformly across all subjects would risk under- or over-amplification of tissue signals in individuals with atypical acoustic properties (e.g., variation in buccal fat thickness, mucosal density, or submucosal tissue composition), producing images of reduced diagnostic quality that would not reflect realistic clinical acquisition conditions.

The distribution of adjustable parameters across the final 40-subject cohort is summarized in Table S1. For each subject, parameters were set at the beginning of the examination and kept consistent across scanning positions wherever possible; intra-subject gain adjustments occurred in 13 of 40 subjects (per-subject ranges in Table S3).

**Table S1.** Acquisition parameter distribution across the final 40-subject cohort (477 images).

| Parameter | Value or distribution |
|---|---|
| Operating frequency | 11.5 MHz (all subjects) |
| Acoustic power output | 100% (all subjects) |
| Imaging preset | Superficial 2D B-mode: 37 subjects (442 images); Thyroid 2D B-mode: 3 subjects (35 images) |
| Dynamic range | 80 dB: 37 subjects (442 images); 78 dB: 3 subjects (35 images) |
| Gain | Range 53–73 dB; median 65 dB; mean 64.8 ± 5.0 dB |

| Parameter | Value or distribution |
|---|---|
| Imaging depth | Range 2.0–2.75 cm; median 2.75 cm |
| Intra-subject gain consistency | Fully constant across all images: 27/40 subjects; Intra-subject variation: 13/40 subjects |

*The three subjects imaged with the Thyroid preset (IO4, V5, V29) were allocated to the training/validation set; all 8 test-set subjects were imaged with the Superficial preset (see Section S3.3).*

##### S2. Dataset Origin and Image Exclusion

The intraoral ultrasound dataset analysed in this study was originally acquired for anatomical investigation of the buccal mucosa, prior to the conception of the present deep learning analysis. Imaging was therefore performed under routine clinical acquisition conditions rather than a research protocol prospectively designed for deep learning. Approval by the Institutional Review Board of Yonsei University Dental Hospital (approval no. 2-2023-0050) covered both the primary anatomical research and the present secondary computational analysis.

A total of 531 intraoral ultrasound images were originally acquired from the 40 healthy volunteers. Prior to any model training or analysis, 54 images (10.2%) were excluded following quality assessment by a single trained operator using the following criteria:

(a) File corruption: images exhibiting data loss or rendering artefacts (e.g., partial image frames, grey replacement blocks) introduced during file transfer or archival, rendering the image unusable.

(b) Inadequate anatomical visualization: images in which poor acoustic coupling, probe contact failure, or excessive attenuation resulted in incomplete visualization of the target tissue layers (dermis, submucosa, and muscle), such that anatomical structures required for class identification could not be reliably resolved.

(c) Modality mismatch: colour Doppler overlay images acquired for vascular assessment during the same examination sessions, which contained hue-encoded velocity information incompatible with the greyscale B-mode classification task of the present study.

Exclusion was conducted prior to subject-level partitioning and independently of any class label or classification procedure. Excluded images were distributed across 25 of 40 subjects (mean 2.2 excluded images per affected subject; range 1–6). Exclusion rates differed modestly between classes (Inferior: 37/274, 13.5%; Superior: 17/257, 6.6%), primarily reflecting a higher rate of technical acquisition issues in the most posterior inferior region (IBM3: 18 excluded), which corresponds to the anatomically least accessible area of the buccal mucosa for intraoral probe placement. Despite this differential exclusion, the final analytic dataset maintained near-perfect class balance (Inferior: 237 images, 49.7%; Superior: 240 images, 50.3%), and exclusion preceded any classification analysis.

##### S3. Acquisition parameters by class

Acquisition parameters were selected per subject. All 40 subjects contributed both classes in one session, reducing differences in subject-level preferences between classes. The analyses below describe the measured parameter distributions; non-significant tests do not establish independence or rule out unmeasured confounding.

###### S3.1 Image-Level Parameter Comparisons (n = 477)

At the image level, no significant association was observed between any acquisition parameter and class label (Table S2). Mean gain was effectively identical between classes (Inferior: 64.78 ± 4.93 dB; Superior: 64.73 ± 5.12 dB; Mann–Whitney U = 28343.5, p = 0.949). Preset distribution was balanced (Inferior: 17 Thyroid / 220 Superficial; Superior: 18 Thyroid / 222 Superficial; Fisher's exact test, p = 1.000). Mean imaging depth was identical to three decimal places between classes (both 2.706 cm; Mann–Whitney U = 28407.0, p = 0.966).

##### S3.2 Within-Subject Comparison

Each of the 40 subjects contributed both Inferior and Superior images; consequently, any subject-level baseline acquisition preference is automatically matched between classes within that subject. Of the 13 subjects in whom any intra-subject gain variation occurred, 12 exhibited a non-zero mean gain difference between Inferior and Superior acquisitions. However, the direction of this within-subject variation was not systematic: the mean paired difference (Superior minus Inferior) across all 40 subjects was −0.036 dB, the paired Wilcoxon signed-rank test (n=12, excluding 28 zero-difference pairs) did not detect class-directional asymmetry (W = 34.5, p = 0.750).

##### S3.3 Test Set Parameter Uniformity

Because all Grad-CAM interpretability analyses reported in the main text were conducted on the held-out test set (n = 97, 8 subjects), the confound structure of this specific subset is of particular importance. The test set exhibited greater parameter uniformity than the full dataset: all 97 test images were acquired using the Superficial 2D B-mode preset with dynamic range 80 dB (the three Thyroid-preset subjects were allocated to training/validation only). Within the test set, gain values had similar observed distributions between classes (Inferior: 63.62 ± 5.26 dB; Superior: 63.61 ± 5.51 dB; Mann–Whitney U = 1162.5, p = 0.925), and imaging depth was restricted to 2.5–2.75 cm with no difference detected by the exploratory image-level test (Mann–Whitney U p = 0.975).

##### S3.4 Implication for Interpretability Analyses

The measured parameter distributions were similar between classes, and the paired subject-level gain comparison did not detect a directional difference. These observations reduce concern about the measured variables but do not exclude acquisition confounding or identify the features responsible for the Grad-CAM patterns. Image-level tests do not account for within-subject dependence.

Table S2. Acquisition parameter comparisons by class: statistical test summary.

| Parameter | Analysis level | Test | Inferior | Superior | p-value |
|---|---|---|---|---|---|
| Gain | Image (n = 477) | Mann–Whitney U | 64.78 ± 4.93 dB (n = 237) | 64.73 ± 5.12 dB (n = 240) | 0.949 |
| Preset | Image (n = 477) | Fisher's exact | 17 Thy / 220 Sup | 18 Thy / 222 Sup | 1.000 |
| Depth | Image (n = 477) | Mann–Whitney U | 2.706 ± 0.158 cm | 2.706 ± 0.157 cm | 0.966 |
| Gain (paired) | Subject (n = 40) | Wilcoxon signed-rank | Paired difference (Sup − Inf): mean −0.036 dB | | 0.750 |
| Gain | Test set (n = 97) | Mann–Whitney U | 63.62 ± 5.26 dB (n = 48) | 63.61 ± 5.51 dB (n = 49) | 0.925 |
| Depth | Test set (n = 97) | Mann–Whitney U | 2.719 ± 0.084 cm | 2.719 ± 0.083 cm | 0.975 |
| Preset | Test set (n = 97) | — | All Superficial (n = 48) | All Superficial (n = 49) | — |

Values for gain and depth are mean ± SD. All tests are two-sided. Image-level tests are exploratory and do not account for subject clustering. The Wilcoxon signed-rank test on paired subject-level gain differences excluded 28 subjects with zero paired between-class mean gain difference, per standard Wilcoxon Convention.

**Table S3.** Per-subject acquisition parameters for the final 40-subject cohort (477 images).

| Subject | Split | N images | Preset | DR (dB) | Gain (dB) | Depth (cm) |
|---|---|---|---|---|---|---|
| IO2 | Train/Val | 11 | Superficial | 80 | 68 | 2 |
| IO3 | Train/Val | 12 | Superficial | 80 | 68 | 2.75 |
| IO4 | Train/Val | 11 | Thyroid | 78 | 66 | 2.75 |
| IO5 | Train/Val | 13 | Superficial | 80 | 53–63 (mean: 61.8) | 2.75 |
| V5 | Train/Val | 12 | Thyroid | 78 | 73 | 2.75 |
| V6 | Train/Val | 12 | Superficial | 80 | 53 | 2.75 |
| V7 | Test | 12 | Superficial | 80 | 53–56 (mean: 53.8) | 2.5 |
| V8 | Train/Val | 12 | Superficial | 80 | 58 | 2.5 |
| V9 | Train/Val | 12 | Superficial | 80 | 66 | 2.75 |
| V10 | Test | 12 | Superficial | 80 | 61 | 2.75 |
| V11 | Train/Val | 12 | Superficial | 80 | 65 | 2.75 |
| V12 | Train/Val | 12 | Superficial | 80 | 56–65 (mean: 64.0) | 2.75 |
| V13 | Train/Val | 12 | Superficial | 80 | 53–58 (mean: 57.6) | 2.75 |
| V14 | Train/Val | 12 | Superficial | 80 | 68 | 2.75 |
| V15 | Train/Val | 12 | Superficial | 80 | 60 | 2.75 |
| V16 | Train/Val | 12 | Superficial | 80 | 59 | 2.75 |
| V17 | Train/Val | 9 | Superficial | 80 | 59–62 (mean: 61.7) | 2 |
| V18 | Test | 12 | Superficial | 80 | 64 | 2.75 |
| V19 | Train/Val | 12 | Superficial | 80 | 65 | 2.75 |
| V20 | Train/Val | 12 | Superficial | 80 | 57–60 (mean: 59.8) | 2.75 |
| V21 | Test | 12 | Superficial | 80 | 69 | 2.75 |
| V22 | Test | 13 | Superficial | 80 | 68 | 2.75 |
| V23 | Train/Val | 12 | Superficial | 80 | 67 | 2.75 |
| V24 | Train/Val | 12 | Superficial | 80 | 71 | 2.75 |
| V25 | Test | 12 | Superficial | 80 | 71 | 2.75 |
| V26 | Train/Val | 12 | Superficial | 80 | 65 | 2.75 |
| V27 | Train/Val | 12 | Superficial | 80 | 69 | 2.75 |
| V28 | Train/Val | 12 | Superficial | 80 | 62–68 (mean: 63.0) | 2.75 |
| V29 | Train/Val | 12 | Thyroid | 78 | 72 | 2.75 |

| Subject | Split | N images | Preset | DR (dB) | Gain (dB) | Depth (cm) |
|---|---|---|---|---|---|---|
| V30 | Train/Val | 12 | Superficial | 80 | 59–68 (mean: 60.8) | 2.75 |
| V31 | Train/Val | 12 | Superficial | 80 | 62 | 2.75 |
| V32 | Test | 12 | Superficial | 80 | 58–62 (mean: 60.2) | 2.75 |
| V33 | Test | 12 | Superficial | 80 | 58–62 (mean: 61.7) | 2.75 |
| V34 | Train/Val | 12 | Superficial | 80 | 65 | 2.75 |
| V35 | Train/Val | 12 | Superficial | 80 | 65–70 (mean: 69.6) | 2.75 |
| V36 | Train/Val | 12 | Superficial | 80 | 67–70 (mean: 68.5) | 2.75 |
| V37 | Train/Val | 12 | Superficial | 80 | 66 | 2.75 |
| V38 | Train/Val | 12 | Superficial | 80 | 71 | 2.75 |
| V39 | Train/Val | 12 | Superficial | 80 | 63–67 (mean: 64.7) | 2.75 |
| V40 | Train/Val | 12 | Superficial | 80 | 73 | 2.75 |

Subjects are identified by anonymized codes. "Gain (dB)" is reported as a single value when constant across all images of the subject, or as "min–max (mean: X)" when intra-subject variation occurred during acquisition. All 40 subjects contributed both Superior and Inferior images (Inferior: 237 images; Superior: 240 images). The three subjects imaged with the Thyroid preset (IO4, V5, V29) were allocated to the Training/Validation set; all 8 Test set subjects were imaged with the Superficial preset. Per-subject summaries are provided here; per-image acquisition records are available from the corresponding author on reasonable request.

##### S4. Hyperparameter Optimization Results

Hyperparameters were optimized independently with Optuna and Tree-structured Parzen Estimator sampling, using 20 trials per architecture. ResNet-50, DenseNet-121 and EfficientNet were tuned on Fold 1; ConvNeXt-Tiny and DeiT-Tiny were tuned on Fold 2. Table S4 reports the settings for all five architectures. Search spaces are specified in main-text Section 2.5; single-fold tuning and differences between training pipelines are discussed in Section 4.7.

**Table S4. Optimal hyperparameters for ResNet-50, DenseNet-121 and EfficientNet-B0 (20 Optuna trials per architecture; Fold 1 tuning). ConvNeXt-Tiny and DeiT-Tiny were tuned on Fold 2. ConvNeXt-Tiny selected learning rate 2.36886395 × $10^{-5}$, Adam, batch size 8 and maximum 21 epochs; DeiT-Tiny selected learning rate 4.32845022 × $10^{-6}$, AdamW (weight decay 0.05), batch size 8 and maximum 24 epochs. Search spaces are specified in main-text Section 2.5.**

| Hyperparameter | Search Space | Optimal: ResNet-50 | Optimal: DenseNet-121 | Optimal: EfficientNet-B0 |
|---|---|---|---|---|
| Learning rate | $1\times10^{-5}$ to $1\times10^{-4}$ (log) | $5.24\times10^{-5}$ | $7.35\times10^{-5}$ | $1.87\times10^{-5}$ |
| Batch Size | {8, 16, 32} | 8 | 16 | 16 |
| Optimizer | {Adam, SGD} | Adam | Adam | Adam |

| Hyperparameter | Search Space | Optimal: ResNet-50 | Optimal: DenseNet-121 | Optimal: EfficientNet-B0 |
|---|---|---|---|---|
| Max Epochs | 20 to 50 | 20 | 26 | 27 |

#### S5. Cross-Validation Performance Across Folds

Five-fold subject-independent cross-validation (GroupKFold) was used to assess generalization stability. Each fold contained approximately 6–7 subjects with no patient overlap. Table S5 reports per-fold F1-scores and the mean ± standard deviation across folds. The CV F1–Test F1 difference (Δ) is reported descriptively.

| Architecture | Fold 1 | Fold 2 | Fold 3 | Fold 4 | Fold 5 | CV Mean ± SD | Test F1 | ΔF1 |
|---|---|---|---|---|---|---|---|---|
| ResNet-50 | 0.841 | 1.000 | 0.900 | 0.850 | 0.886 | 0.895 ± 0.063 | 0.874 | −0.021 |
| DenseNet-121 | 0.785 | 0.935 | 0.886 | 0.853 | 0.875 | 0.867 ± 0.055 | 0.911 | +0.044 |
| EfficientNet-B0 | 0.804 | 0.925 | 0.907 | 0.800 | 0.850 | 0.857 ± 0.058 | 0.929 | +0.072 |
| EfficientNet-B7 (excluded) | 0.554 | 0.610 | 0.679 | 0.645 | 0.660 | 0.630 ± 0.049 | Excluded | — |

Table S5. Five-fold subject-independent cross-validation results (GroupKFold) for the four evaluated architectures. Per-fold F1-scores, cross-validation mean ± SD, held-out test F1 and their descriptive difference ΔF1 (Test − CV) are reported. EfficientNet-B7 was excluded from final evaluation owing to its lower cross-validation performance; overparameterization was not tested directly.

#### S6. Layer-Wise Grad-CAM Target Layer Selection

Target layers for Grad-CAM analysis were selected through empirical visual inspection of activation patterns across multiple candidate layers within ResNet-50, DenseNet-121 and EfficientNet-B0. Although the deepest available layer typically exhibited the highest mean activation intensity, activations in those layers were spatially concentrated in broad, anatomically non-specific regions. Intermediate layers appeared on visual inspection to show more coherent patterns near tissue boundaries while preserving spatial resolution; anatomical correspondence was not quantitatively validated. Figure S1 illustrates this pattern for ResNet-50. ConvNeXt-Tiny used model.features[−1], and the non-distilled DeiT-Tiny model used model.blocks[−1].norm1. For DeiT-Tiny, the class token was discarded and the 196 patch tokens were reshaped to 14 × 14 and transposed to channel-first format. These choices were also used for the alternative CAM methods; no candidate-layer comparison was performed for these two models in the archived implementation.

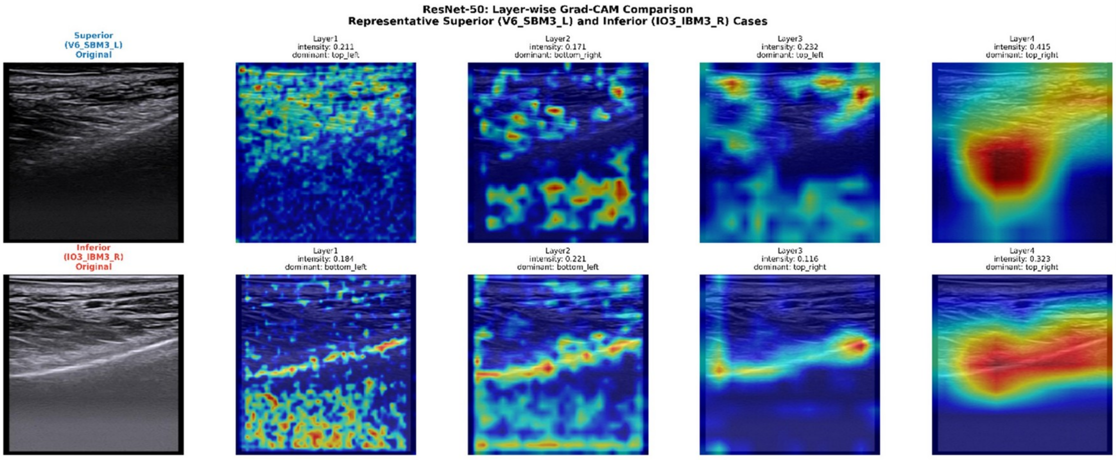


**Figure S1.** *Layer-wise Grad-CAM comparison for representative Superior (V6_SBM3_L) and Inferior (IO3_IBM3_R) cases of ResNet-50. Top row: Superior case original ultrasound image followed by Layer1–Layer4 Grad-CAM heatmaps. Bottom row: Inferior case original image followed by Layer1–Layer4 heatmaps. Mean activation intensity and dominant activation quadrant are annotated for each layer. Although Layer4 exhibited the highest mean intensity (Superior 0.415; Inferior 0.323), activations were concentrated in broad, anatomically non-specific regions. On visual inspection, Layer3 appeared to show more coherent patterns near tissue boundaries; in the Inferior example, a linear activation pattern appeared near the hyperechoic dermis line (this single-run visual observation informed target-layer selection only; it is not evidence of anatomical alignment, see main-text Section 4.5 and Supplementary Table S7). ResNet-50 is shown as a representative example; analogous visual inspection across candidate layers informed the corresponding target-layer selections for DenseNet-121 (DenseBlock3) and EfficientNet-B0 (features[−3]).*

#### S7. Performance vs. Computational Efficiency

Figure S2 visualizes the relationship between classification performance (F1-score) and per-image inference latency across the three retained architectures, with marker size proportional to parameter count.

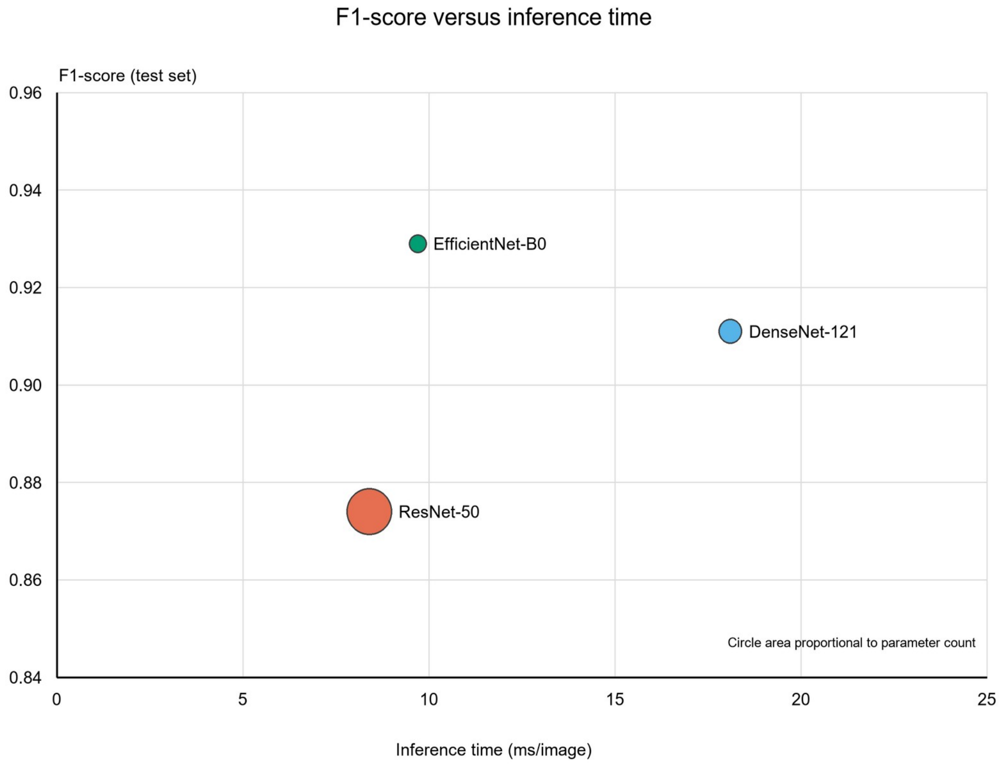


Figure S2. Classification performance (F1-score) versus per-image inference time for the original best-fold ResNet-50, DenseNet-121 and EfficientNet-B0 models. Marker size is proportional to parameter count. This descriptive figure does not establish architectural superiority or equivalence.

#### S8. Extended Misclassification Analysis

Figures S3 and S4 provide descriptive examples and spatial-metric comparisons for the three original best-fold models (main text Sections 3.7 and 4.6). The exploratory image-level comparisons do not account for subject clustering and cannot establish that ambiguity caused the errors or that attention strategies are equivalent.

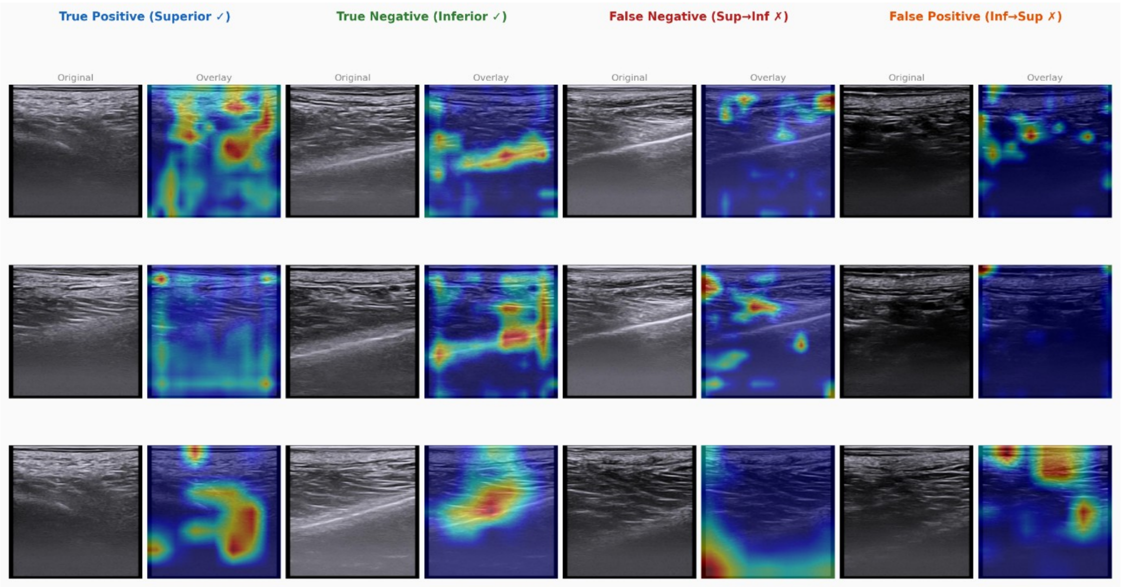


**Figure S3.** *Extended misclassification gallery: representative true positive (Superior ✓), true negative (Inferior ✓), false negative (Superior → Inferior ✗), and false positive (Inferior → Superior ✗) cases for each of the three retained architectures. Original ultrasound images are shown alongside their Grad-CAM overlays. Selected misclassified examples showed indistinct tissue boundaries; these observations do not establish the cause of misclassification.*

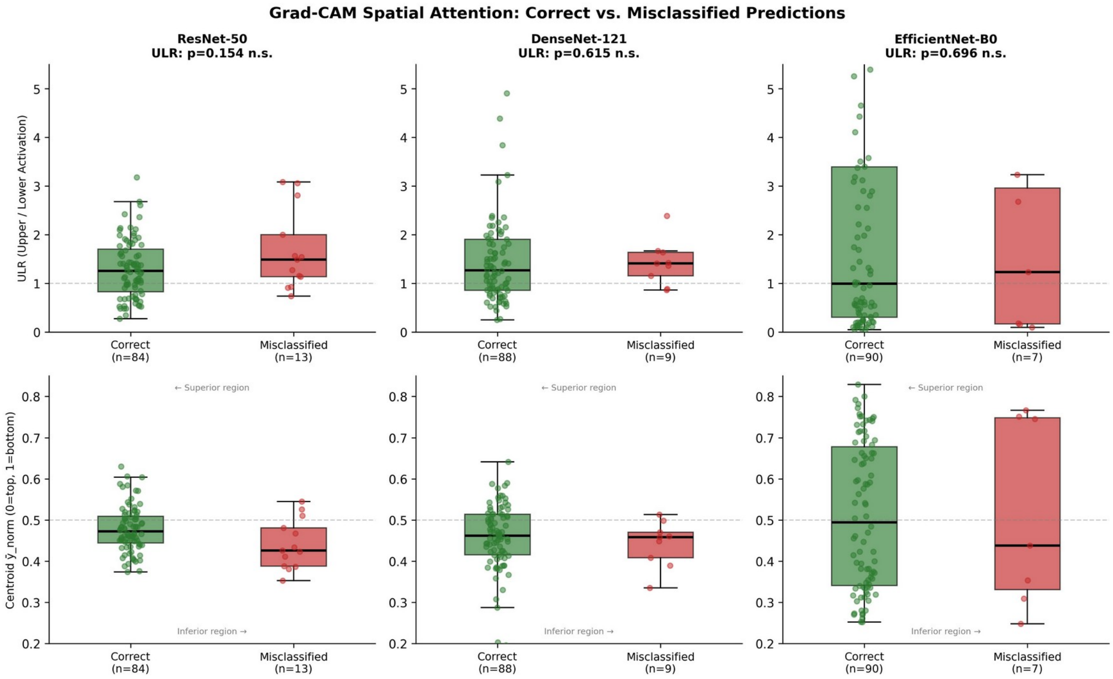


Figure S4. Grad-CAM spatial attention metrics for correctly classified versus misclassified test images, by architecture. Top row: upper-to-lower activation ratio (ULR). Bottom row: normalized vertical centroid $\bar{y}$_norm (0 = image top, 1 = image bottom). Exploratory image-level between-group comparisons, unadjusted for subject clustering, by Mann–Whitney U test (two-sided); no significant differences in ULR were observed for any model (ResNet-50 $p = 0.154$; DenseNet-121 $p = 0.615$; EfficientNet-B0 $p = 0.696$). Sample sizes: ResNet-50 (correct n = 84, misclassified n = 13); DenseNet-121 (88, 9); EfficientNet-B0 (90, 7).

**S9. Label Validation Responses**

Table S6. Per-image expert re-classification responses in the label-validation sub-study (Sections 2.10 and 3.2 of the main text). Image codes are anonymized. Examiner 1 (E1) is the acquiring operator (intra-rater / label-grounding); Examiner 2 (E2) is an independent anatomy expert. Acquisition label = class defined by probe position at acquisition. Confidence rated 1–5. Rows marked * for E1 are re-presented duplicates used to assess intra-rater repeatability (5 pairs) and are therefore additional to the 20 unique images.

| Examiner | Image code | Acquisition label | Expert class | Confidence | Agree |
|---|---|---|---|---|---|
| E1 | img_001 | Sup | Sup | 5 | Yes |
| E1 | img_002 | Sup | Sup | 5 | Yes |
| E1 | img_003 | Inf | Inf | 4 | Yes |
| E1 | img_004 * | Inf | Inf | 5 | Yes |
| E1 | img_005 | Inf | Inf | 5 | Yes |
| E1 | img_006 | Inf | Inf | 3 | Yes |
| E1 | img_007 | Sup | Sup | 4 | Yes |
| E1 | img_008 | Inf | Inf | 5 | Yes |
| E1 | img_009 * | Inf | Inf | 5 | Yes |
| E1 | img_010 * | Sup | Sup | 4 | Yes |
| E1 | img_011 | Sup | Sup | 3 | Yes |
| E1 | img_012 | Sup | Sup | 4 | Yes |
| E1 | img_013 | Sup | Inf | 3 | No |
| E1 | img_014 | Inf | Inf | 5 | Yes |
| E1 | img_015 | Inf | Sup | 3 | No |
| E1 | img_016 | Inf | Inf | 4 | Yes |
| E1 | img_017 | Inf | Inf | 4 | Yes |
| E1 | img_018 | Inf | Inf | 3 | Yes |
| E1 | img_019 | Sup | Sup | 4 | Yes |
| E1 | img_020 | Sup | Sup | 4 | Yes |
| E1 | img_021 | Sup | Sup | 2 | Yes |
| E1 | img_022 * | Inf | Inf | 3 | Yes |
| E1 | img_023 * | Inf | Inf | 3 | Yes |
| E1 | img_024 | Inf | Inf | 4 | Yes |
| E1 | img_025 | Sup | Sup | 5 | Yes |
| E2 (independent) | e2_001 | Sup | Sup | 4 | Yes |

| Examiner | Image code | Acquisition label | Expert class | Confidence | Agree |
|---|---|---|---|---|---|
| E2 (independent) | e2_002 | Inf | Inf | 5 | Yes |
| E2 (independent) | e2_003 | Inf | Inf | 4 | Yes |
| E2 (independent) | e2_004 | Sup | Sup | 5 | Yes |
| E2 (independent) | e2_005 | Inf | Inf | 3 | Yes |
| E2 (independent) | e2_006 | Inf | Inf | 5 | Yes |
| E2 (independent) | e2_007 | Inf | Inf | 4 | Yes |
| E2 (independent) | e2_008 | Inf | Inf | 3 | Yes |
| E2 (independent) | e2_009 | Inf | Inf | 5 | Yes |
| E2 (independent) | e2_010 | Sup | Inf | 3 | No |
| E2 (independent) | e2_011 | Sup | Sup | 5 | Yes |
| E2 (independent) | e2_012 | Inf | Sup | 3 | No |
| E2 (independent) | e2_013 | Inf | Inf | 5 | Yes |
| E2 (independent) | e2_014 | Sup | Inf | 3 | No |
| E2 (independent) | e2_015 | Sup | Sup | 5 | Yes |
| E2 (independent) | e2_016 | Sup | Sup | 5 | Yes |
| E2 (independent) | e2_017 | Sup | Sup | 4 | Yes |
| E2 (independent) | e2_018 | Inf | Inf | 5 | Yes |
| E2 (independent) | e2_019 | Sup | Sup | 5 | Yes |
| E2 (independent) | e2_020 | Sup | Sup | 4 | Yes |
| E2 (independent) | e2_021 | Inf | Inf | 3 | Yes |
| E2 (independent) | e2_022 | Inf | Inf | 5 | Yes |
| E2 (independent) | e2_023 | Inf | Sup | 2 | No |
| E2 (independent) | e2_024 | Inf | Inf | 5 | Yes |
| E2 (independent) | e2_025 | Sup | Sup | 5 | Yes |
| E2 (independent) | e2_026 | Inf | Inf | 4 | Yes |
| E2 (independent) | e2_027 | Sup | Inf | 4 | No |
| E2 (independent) | e2_028 | Sup | Sup | 4 | Yes |
| E2 (independent) | e2_029 | Sup | Sup | 5 | Yes |
| E2 (independent) | e2_030 | Inf | Inf | 5 | Yes |
| E2 (independent) | e2_031 | Sup | Sup | 5 | Yes |
| E2 (independent) | e2_032 | Sup | Inf | 2 | No |

| Examiner | Image code | Acquisition label | Expert class | Confidence | Agree |
|---|---|---|---|---|---|
| E2 (independent) | e2_033 | Inf | Inf | 4 | Yes |
| E2 (independent) | e2_034 | Inf | Inf | 4 | Yes |
| E2 (independent) | e2_035 | Inf | Inf | 5 | Yes |
| E2 (independent) | e2_036 | Sup | Sup | 3 | Yes |
| E2 (independent) | e2_037 | Inf | Inf | 5 | Yes |
| E2 (independent) | e2_038 | Sup | Sup | 4 | Yes |
| E2 (independent) | e2_039 | Sup | Sup | 5 | Yes |
| E2 (independent) | e2_040 | Sup | Sup | 3 | Yes |

##### S10. Robustness, Alternative CAM, and Calibration Analyses

Table S7. Grad-CAM spatial metrics across five seeds. "Same direction" is the largest number of all five seeds sharing the direction of the between-class mean difference, irrespective of significance. "Exploratory P<0.05" counts seeds with an image-level Mann–Whitney result below 0.05; those tests do not account for subject clustering. |r| is the absolute rank-biserial effect size. All three metrics have majority-direction counts of 3/5 for ResNet-50, DenseNet-121, EfficientNet-B0 and DeiT-Tiny, and 4/5 for ConvNeXt-Tiny. These are descriptive counts, not a validated stability threshold.

| Architecture | Metric | Exploratory P<0.05 (/5) | Same direction (/5) | \|r\| range |
|---|---|---|---|---|
| ResNet-50 | intensity | 3 | 3 | 0.07–0.81 |
| | centroid | 3 | 3 | 0.17–0.61 |
| | ULR | 3 | 3 | 0.05–0.62 |
| DenseNet-121 | intensity | 2 | 3 | 0.12–0.94 |
| | centroid | 2 | 3 | 0.04–0.63 |
| | ULR | 3 | 3 | 0.07–0.70 |
| EfficientNet-B0 | intensity | 2 | 3 | 0.08–0.46 |
| | centroid | 4 | 3 | 0.02–0.86 |
| | ULR | 4 | 3 | 0.14–0.78 |
| ConvNeXt-Tiny | intensity | 4 | 4 | 0.08–0.77 |
| | centroid | 4 | 4 | 0.12–0.79 |
| | ULR | 4 | 4 | 0.12–0.73 |
| DeiT-Tiny | intensity | 5 | 3 | 0.44–0.94 |
| | centroid | 4 | 3 | 0.03–0.91 |
| | ULR | 4 | 3 | 0.11–0.85 |

Table S8. Between-class Grad-CAM comparisons under alternative CAM methods (Grad-CAM++, Score-CAM, Layer-CAM). Direction = class with the higher value (Sup>Inf or Inf>Sup); p = exploratory image-level two-sided Mann–Whitney U, unadjusted for subject clustering. The primary single-run Grad-CAM pattern is not consistently reproduced: ResNet-50's ULR significance is lost, and EfficientNet-B0's centroid/ULR "inversion" reverses direction under Score-CAM. Intensity was not computed for ConvNeXt-Tiny and DeiT-Tiny.

| Architecture | Method | Centroid dir (p) | ULR dir (p) | Intensity dir (p) |
|---|---|---|---|---|
| ResNet-50 | Grad-CAM++ | Sup>Inf (0.95) | Sup>Inf (0.10) | Sup>Inf (0.14) |
| | Score-CAM | Sup>Inf (1.8e-10) | Sup>Inf (0.19) | Sup>Inf (6.1e-10) |
| | Layer-CAM | Sup>Inf (0.17) | Sup>Inf (0.66) | Sup>Inf (0.020) |
| DenseNet-121 | Grad-CAM++ | Inf>Sup (0.056) | Sup>Inf (0.068) | Sup>Inf (0.86) |
| | Score-CAM | Inf>Sup (0.051) | Sup>Inf (0.008) | Inf>Sup (2.1e-4) |
| | Layer-CAM | Sup>Inf (0.027) | Inf>Sup (0.17) | Inf>Sup (0.23) |
| EfficientNet-B0 | Grad-CAM++ | Sup>Inf (2.9e-15) | Inf>Sup (1.4e-15) | Sup>Inf (0.23) |
| | Score-CAM | Inf>Sup (2.1e-6) | Sup>Inf (1.9e-6) | Inf>Sup (0.91) |
| | Layer-CAM | Sup>Inf (5.2e-16) | Inf>Sup (1.1e-15) | Sup>Inf (1.1e-5) |
| ConvNeXt-Tiny | Grad-CAM++ | Inf>Sup (0.045) | Sup>Inf (0.12) | — |
| | Score-CAM | Inf>Sup (0.12) | Sup>Inf (0.025) | — |
| | Layer-CAM | Inf>Sup (0.055) | Sup>Inf (0.17) | — |
| DeiT-Tiny | Grad-CAM++ | Inf>Sup (0.12) | Sup>Inf (0.019) | — |
| | Score-CAM | Inf>Sup (3.4e-13) | Sup>Inf (3.1e-13) | — |
| | Layer-CAM | Sup>Inf (5.0e-8) | Sup>Inf (1.7e-8) | — |

Table S9. Model calibration on the held-out test set (n = 97) for the three retained architectures. ECE = expected calibration error (15 equal-width confidence bins); Brier = Brier score. Values describe the original best-fold runs, not the follow-up five-seed calibration in main Table 3. ECE is sensitive to bin count (e.g., DenseNet-121 ECE ranges 0.064–0.087 for 6–15 bins); Brier is bin-independent.

| Model | ECE (15-bin) | Brier score |
|---|---|---|
| ResNet-50 | 0.092 | 0.106 |
| DenseNet-121 | 0.087 | 0.071 |
| EfficientNet-B0 | 0.077 | 0.064 |

##### S11. Participants, Eligibility and Sample Size

The images analysed here were acquired in the parent anatomical study (approved by the Institutional Review Board of Yonsei University Dental Hospital as a minimal-risk study; approval no. 2-2023-0050, approved on 19 October 2023). The approved protocol period was 19 October 2023 to 18 October 2025; participant enrolment and imaging took place between 29 July and 29 August 2024.

Volunteers were recruited by a public notice approved by the Institutional Review Board, and those who responded and consented were enrolled; the series was therefore one of convenience rather than a consecutive or randomly sampled series. Eligibility criteria specified in the approved protocol were: age 19 to 60 years; no missing teeth (third molars excepted); no fixed orthodontic appliance or denture; capacity to give voluntary informed consent after explanation of the purpose and procedures; and no invasive procedure or surgery of the temporalis or masseter region within the preceding six months. Exclusion criteria were: history of trauma to the temporomandibular joint; temporomandibular disc displacement; severe malocclusion, including anterior open bite; invasive procedure or surgery of the temporalis or masseter region within six months; and any other condition judged unsuitable by the examining clinician.

The sample size of the parent study was determined a priori (G*Power 3.1.9.7; two-group t test, large effect size 0.75, alpha = 0.05, power = 0.80), yielding 46 participants. That calculation was made for the anatomical endpoints of the parent study and not for any deep learning analysis. No separate sample size was prespecified for the present secondary analysis, which used every image in the existing dataset that passed quality assessment. Forty of the enrolled participants contributed the buccal mucosa image sets analysed here.

Patient identifiers were never entered on the ultrasound system: each examination was recorded under the study code alone (for example V6, IO3), which is the identifier visible in the image banner and the identifier used throughout this work. No name, hospital number, date of birth or other direct identifier appears in any image.

**S12. Occlusion Boundary Sensitivity, Area-Matched Control and Output Diagnostics**

All analyses in this section were performed by inference only, using the stored seed checkpoints; no model was retrained and no training data were touched. Twenty-five models (five architectures × five random seeds) were evaluated on the same held-out test set (n = 97) used throughout.

**Boundary sweep.** The occlusion boundary was placed at 30%, 40%, 50%, 60% and 70% of image height. At each boundary the region above or the region below was replaced with a constant value of 0.0 in the standardized input, corresponding to the training-set mean intensity of 0.3162 before standardization, giving 50 conditions per architecture (five seeds × five boundaries × two directions). Because the two directions are complementary at a given boundary, the fraction of the image removed ranged from 30% to 70% in each direction.

**Area-matched control.** A contiguous band covering 50% of image height was placed at five vertical offsets spanning the available travel — with the band starting at 0%, 25%, 50%, 75% and 100% of the distance between the top and bottom limits — so that the quantity of removed information was held constant while its location varied. The 0% and 100% offsets coincide with upper-region and lower-region occlusion at the 50% boundary and serve as internal consistency checks.

**Output diagnostics.** For each model and condition, the predicted probability of the superior class was recorded for all 97 images, and from these we computed the standard deviation, the interquartile range, the number of distinct values to three decimal places, the expected calibration error (15 equal-width bins), the Brier score, the mean confidence on incorrect predictions, and the proportion of probabilities below 0.01 or above 0.99.

Collapse criterion. Collapse was operationally defined as a held-out AUC below 0.60. Because the threshold is itself a choice, collapse rates were additionally computed at 0.55, 0.65 and 0.70 and are reported in the main text.

Reproducibility note. The original five-seed metrics in main Table 1 are retained from the training-run JSON records. The follow-up sensitivity execution used saved checkpoints in a different computing environment; the inference batch size also changed from 64 to 97 images. Baseline AUC agreed within 0.002 for all models, and masked AUC within 0.003 for four architectures. EfficientNet-B0 masked AUC differed by up to 0.161, while baseline differed by less than 0.001. The relative contributions of environment, batch size and saturated outputs were not isolated. In the follow-up environment, two repeated evaluations of baseline and upper 50% occlusion gave identical probabilities (maximum absolute difference 0, Spearman's ρ approximately 1). This repeatability check did not cover every boundary or lower-mask condition and does not explain differences between executions. Main Table 3 and Supplementary Table S14 use follow-up results; the latter's intervals must not be applied to original Table 1 values.

Table S10. Classification performance and computational efficiency on the held-out test set (n = 97) for the single best-fold run of the three architectures carried through the detailed analyses. Ninety-five per cent confidence intervals were estimated by image-level bootstrap resampling (n = 1,000), unadjusted for subject clustering and interpreted as exploratory using the same test-set predictions as the occlusion ablation analysis (Table 2 of the main text). TN = true negative; FP = false positive; FN = false negative; TP = true positive. The primary analyses in the main text are seed-averaged (Table 1); this table is provided for the condition-wise comparisons that use the same single run.

| **Model** | **Params (M)** | **F1-Score (95% CI)** | **Precision (95% CI)** | **Recall (95% CI)** | **AUC (95% CI)** | **Inference (ms/image)** | **TN/FP/FN/TP** |
|---|---|---|---|---|---|---|---|
| Resnet-50 | 25.6 | 0.874 [0.796–0.932] | 0.833 [0.732–0.927] | 0.918 [0.840–0.982] | 0.945 [0.895–0.981] | 8.4 ± 3.2 | 39 / 9 / 4 / 45 |
| DenseNet-121 | 7.0 | 0.911 [0.844–0.962] | 0.885 [0.796–0.964] | 0.939 [0.867–1.000] | 0.963 [0.926–0.990] | 18.1 ± 0.5 | 42 / 6 / 3 / 46 |
| EfficientNet-B0 | 4.0 | 0.929 [0.870–0.975] | 0.920 [0.840–0.980] | 0.939 [0.860–1.000] | 0.981 [0.951–0.998] | 9.7 ± 0.8 | 44 / 4 / 3 / 46 |
| EfficientNet-B7 (excluded) | 63.8 | CV: 0.630 ± 0.049 | — | — | CV: 0.793 ± 0.154 | — | — |

Table S11. Label validation via blinded expert re-classification. High confidence = examiner confidence ≥ 4 on a 1–5 scale. Both examiners classified images blinded to the acquisition position and to each other's responses. Examiner 1 acquired the images and defined the acquisition labels, so that comparison is an intra-rater, label-grounding check rather than an independent inter-observer test. κ = Cohen's κ; CI = original image-bootstrap confidence interval, unadjusted for subject clustering; these intervals are exploratory. Per-image responses are in Table S6.

| **Comparison** | **n** | **Agreement (%)** | **Cohen's κ (95% CI)** |
|---|---|---|---|
| Examiner 1, all unique (label-grounding) | 20 | 90.0 | 0.80 [0.49–1.00] |
| — High-confidence (≥4) | 14 | 100 | 1.00 [1.00–1.00] |
| — Intra-rater repeatability (duplicated pairs) | 5 pairs | 100 (5/5) | — |
| Independent expert 2, all | 40 | 85.0 | 0.70 [0.46–0.90] |
| — High-confidence (≥4) | 30 | 96.7 | 0.93 [0.79–1.00] |
| Inter-observer (overlapping images) | 20 | 100 | 1.00 [1.00–1.00] |
| — High-confidence (≥4) | 17 | 100 | 1.00 |

**Table S12.** Distribution of predicted probabilities with and without occlusion, averaged across five random seeds. "Distinct values" is the number of unique predicted probabilities to three decimal places among the 97 test images; the SD ratio is the standard deviation of the predicted probability under occlusion divided by its value at baseline. A ratio above 1 with few distinct values and an interquartile range approaching 1.0 indicates output saturated toward 0 and 1 rather than uncertain output.

| Architecture | Distinct values, baseline | Distinct values, upper / lower occlusion | SD ratio, upper / lower | IQR, upper / lower |
|---|---|---|---|---|
| ResNet-50 | 62.0 | 75.0 / 80.6 | 0.64 / 0.82 | 0.40 / 0.68 |
| DenseNet-121 | 77.6 | 67.0 / 90.6 | 0.47 / 0.64 | 0.13 / 0.47 |
| EfficientNet-B0 | 90.6 | 15.0 / 8.2 | 1.35 / 1.34 | 1.000 / 0.998 |
| ConvNeXt-Tiny | 72.4 | 85.8 / 85.0 | 0.83 / 0.75 | 0.71 / 0.54 |
| DeiT-Tiny | 77.4 | 90.8 / 88.8 | 0.64 / 0.74 | 0.47 / 0.57 |

Table S13. Exploratory image-level pairwise comparison of areas under the receiver operating characteristic curve by the DeLong method, for all ten architecture pairs at each of five random seeds (50 comparisons). ΔAUC is architecture 1 minus architecture 2. These tests do not account for subject clustering and do not establish equivalence or population-level differences. P values were corrected for multiplicity within each seed by the Holm–Bonferroni procedure. Four comparisons reached $P < 0.05$ before correction and one after (ResNet-50 versus ConvNeXt-Tiny at seed 1); the sign of ΔAUC reversed across seeds for all ten pairs, and none of the twenty comparisons involving EfficientNet-B0 reached $P < 0.05$, corrected or uncorrected.

| Seed | Model 1 | Model 2 | AUC 1 | AUC 2 | ΔAUC | z | P | P (Holm) |
|---|---|---|---|---|---|---|---|---|
| 0 | ResNet-50 | DenseNet-121 | 0.9583 | 0.9702 | -0.0119 | -0.610 | 0.5419 | 1.0000 |
| 0 | ResNet-50 | EfficientNet-B0 | 0.9583 | 0.9643 | -0.0060 | -0.325 | 0.7455 | 1.0000 |
| 0 | ResNet-50 | ConvNeXt-Tiny | 0.9583 | 0.9834 | -0.0251 | -1.835 | 0.0665 | 0.6650 |
| 0 | ResNet-50 | DeiT-Tiny | 0.9583 | 0.9685 | -0.0102 | -0.755 | 0.4504 | 1.0000 |
| 0 | DenseNet-121 | EfficientNet-B0 | 0.9702 | 0.9643 | +0.0060 | +0.422 | 0.6728 | 1.0000 |
| 0 | DenseNet-121 | ConvNeXt-Tiny | 0.9702 | 0.9834 | -0.0132 | -1.016 | 0.3098 | 1.0000 |
| 0 | DenseNet-121 | DeiT-Tiny | 0.9702 | 0.9685 | +0.0017 | +0.108 | 0.9142 | 1.0000 |
| 0 | EfficientNet-B0 | ConvNeXt-Tiny | 0.9643 | 0.9834 | -0.0191 | -1.458 | 0.1448 | 1.0000 |
| 0 | EfficientNet-B0 | DeiT-Tiny | 0.9643 | 0.9685 | -0.0043 | -0.242 | 0.8088 | 1.0000 |
| 0 | ConvNeXt-Tiny | DeiT-Tiny | 0.9834 | 0.9685 | +0.0149 | +1.289 | 0.1972 | 1.0000 |
| 1 | ResNet-50 | DenseNet-121 | 0.9133 | 0.9745 | -0.0612 | -2.543 | 0.0110 | 0.0990 |
| 1 | ResNet-50 | EfficientNet-B0 | 0.9133 | 0.9520 | -0.0387 | -1.528 | 0.1265 | 0.8852 |
| 1 | ResNet-50 | ConvNeXt-Tiny | 0.9133 | 0.9685 | -0.0553 | -3.023 | 0.0025 | 0.0250 |
| 1 | ResNet-50 | DeiT-Tiny | 0.9133 | 0.9690 | -0.0557 | -2.483 | 0.0130 | 0.1043 |
| 1 | DenseNet-121 | EfficientNet-B0 | 0.9745 | 0.9520 | +0.0225 | +1.329 | 0.1839 | 1.0000 |
| 1 | DenseNet-121 | ConvNeXt-Tiny | 0.9745 | 0.9685 | +0.0060 | +0.563 | 0.5735 | 1.0000 |

| Seed | Model 1 | Model 2 | AUC 1 | AUC 2 | ΔAUC | z | P | P (Holm) |
|---|---|---|---|---|---|---|---|---|
| 1 | DenseNet-121 | DeiT-Tiny | 0.9745 | 0.9690 | +0.0055 | +0.562 | 0.5738 | 1.0000 |
| 1 | EfficientNet-B0 | ConvNeXt-Tiny | 0.9520 | 0.9685 | -0.0166 | -1.201 | 0.2299 | 1.0000 |
| 1 | EfficientNet-B0 | DeiT-Tiny | 0.9520 | 0.9690 | -0.0170 | -1.275 | 0.2023 | 1.0000 |
| 1 | ConvNeXt-Tiny | DeiT-Tiny | 0.9685 | 0.9690 | -0.0004 | -0.050 | 0.9602 | 1.0000 |
| 2 | ResNet-50 | DenseNet-121 | 0.9741 | 0.9388 | +0.0353 | +1.819 | 0.0690 | 0.6898 |
| 2 | ResNet-50 | EfficientNet-B0 | 0.9741 | 0.9486 | +0.0255 | +1.311 | 0.1899 | 1.0000 |
| 2 | ResNet-50 | ConvNeXt-Tiny | 0.9741 | 0.9626 | +0.0115 | +1.019 | 0.3083 | 1.0000 |
| 2 | ResNet-50 | DeiT-Tiny | 0.9741 | 0.9762 | -0.0021 | -0.202 | 0.8403 | 1.0000 |
| 2 | DenseNet-121 | EfficientNet-B0 | 0.9388 | 0.9486 | -0.0098 | -0.367 | 0.7136 | 1.0000 |
| 2 | DenseNet-121 | ConvNeXt-Tiny | 0.9388 | 0.9626 | -0.0238 | -1.507 | 0.1319 | 0.9231 |
| 2 | DenseNet-121 | DeiT-Tiny | 0.9388 | 0.9762 | -0.0374 | -1.753 | 0.0795 | 0.7158 |
| 2 | EfficientNet-B0 | ConvNeXt-Tiny | 0.9486 | 0.9626 | -0.0140 | -0.708 | 0.4791 | 1.0000 |
| 2 | EfficientNet-B0 | DeiT-Tiny | 0.9486 | 0.9762 | -0.0276 | -1.614 | 0.1065 | 0.8518 |
| 2 | ConvNeXt-Tiny | DeiT-Tiny | 0.9626 | 0.9762 | -0.0136 | -1.192 | 0.2333 | 1.0000 |
| 3 | ResNet-50 | DenseNet-121 | 0.9821 | 0.9354 | +0.0468 | +1.838 | 0.0660 | 0.5944 |
| 3 | ResNet-50 | EfficientNet-B0 | 0.9821 | 0.9694 | +0.0128 | +0.939 | 0.3475 | 1.0000 |
| 3 | ResNet-50 | ConvNeXt-Tiny | 0.9821 | 0.9745 | +0.0077 | +0.716 | 0.4739 | 1.0000 |
| 3 | ResNet-50 | DeiT-Tiny | 0.9821 | 0.9617 | +0.0204 | +1.272 | 0.2034 | 1.0000 |
| 3 | DenseNet-121 | EfficientNet-B0 | 0.9354 | 0.9694 | -0.0340 | -1.681 | 0.0928 | 0.7422 |
| 3 | DenseNet-121 | ConvNeXt-Tiny | 0.9354 | 0.9745 | -0.0391 | -2.028 | 0.0425 | 0.4254 |
| 3 | DenseNet-121 | DeiT-Tiny | 0.9354 | 0.9617 | -0.0264 | -1.029 | 0.3035 | 1.0000 |
| 3 | EfficientNet-B0 | ConvNeXt-Tiny | 0.9694 | 0.9745 | -0.0051 | -0.440 | 0.6603 | 1.0000 |
| 3 | EfficientNet-B0 | DeiT-Tiny | 0.9694 | 0.9617 | +0.0077 | +0.449 | 0.6533 | 1.0000 |
| 3 | ConvNeXt-Tiny | DeiT-Tiny | 0.9745 | 0.9617 | +0.0128 | +0.876 | 0.3811 | 1.0000 |
| 4 | ResNet-50 | DenseNet-121 | 0.9592 | 0.9762 | -0.0170 | -1.072 | 0.2838 | 1.0000 |
| 4 | ResNet-50 | EfficientNet-B0 | 0.9592 | 0.9809 | -0.0217 | -1.372 | 0.1699 | 1.0000 |
| 4 | ResNet-50 | ConvNeXt-Tiny | 0.9592 | 0.9690 | -0.0098 | -0.641 | 0.5217 | 1.0000 |
| 4 | ResNet-50 | DeiT-Tiny | 0.9592 | 0.9494 | +0.0098 | +0.486 | 0.6272 | 1.0000 |
| 4 | DenseNet-121 | EfficientNet-B0 | 0.9762 | 0.9809 | -0.0047 | -0.412 | 0.6805 | 1.0000 |
| 4 | DenseNet-121 | ConvNeXt-Tiny | 0.9762 | 0.9690 | +0.0072 | +0.968 | 0.3330 | 1.0000 |
| 4 | DenseNet-121 | DeiT-Tiny | 0.9762 | 0.9494 | +0.0268 | +1.601 | 0.1094 | 0.9843 |
| 4 | EfficientNet-B0 | ConvNeXt-Tiny | 0.9809 | 0.9690 | +0.0119 | +0.943 | 0.3458 | 1.0000 |
| 4 | EfficientNet-B0 | DeiT-Tiny | 0.9809 | 0.9494 | +0.0315 | +1.922 | 0.0546 | 0.5465 |

| Seed | Model 1 | Model 2 | AUC 1 | AUC 2 | ΔAUC | z | P | P (Holm) |
|---|---|---|---|---|---|---|---|---|
| 4 | ConvNeXt-Tiny | DeiT-Tiny | 0.9690 | 0.9494 | +0.0196 | +1.264 | 0.2062 | 1.0000 |

**Figure S5.** Distribution of predicted probabilities with and without occlusion. Empirical cumulative distribution of the predicted probability of the superior class across the 97 held-out images, shown separately for each architecture and each of five random seeds, without occlusion and under 50% upper-region occlusion. Under occlusion, EfficientNet-B0's predicted probabilities concentrate near 0 and 1, whereas the other four architectures retain a more graded distribution. The figure is shown at the end of this document.

Figure S6. Flow of participants and images. Of 531 images acquired from 40 volunteers, 54 were excluded before partitioning, leaving 477 images. Subject-level partitioning produced 380 training/validation images from 32 subjects and 97 test images from eight subjects. The reference labels for model evaluation were the scanning positions recorded at acquisition. Blinded expert re-classification was a separate label-validation sub-study on selected images (Table S11), not the reference standard for all test images. Recruitment and eligibility are described in Supplementary Methods S11. The figure is shown at the end of this document.

##### S13. Subject-cluster uncertainty estimates

Saved follow-up probabilities were analysed for five architectures, five fixed training seeds and three conditions (baseline, upper 50% and lower 50% occlusion). The 97 images comprised 49 superior and 48 inferior images from eight subjects. In 20,000 paired bootstrap resamples (random seed 20261004), eight subjects were sampled with replacement and all their images were retained with multiplicity. AUC used half credit for exact probability ties; F1 used a superior-class probability threshold of 0.5. Metrics were calculated separately for each fixed trained seed and averaged. The same resamples were used across models and conditions. Percentile 95% intervals describe test-subject uncertainty conditional on these models and predictions. They do not include training-set uncertainty; eight clusters remain a substantial limitation. Per-image probabilities were not available for every boundary-sweep condition, so clustered intervals are limited to the three saved conditions.

Table S14. Follow-up five-seed mean performance with paired subject-cluster 95% confidence intervals. Panel A reports AUC; panel B reports F1; panel C reports baseline AUC minus the average of upper/lower 50% occlusion AUC. These estimates belong to the follow-up execution and differ from original Table 1 values as described in Methods S12. The reduction intervals are within-model comparisons, not tests of differences between architectures.

Panel A. AUC

| Architecture | Baseline | Upper 50% | Lower 50% |
|---|---|---|---|
| ResNet-50 | 0.957 [0.919–0.991] | 0.885 [0.841–0.929] | 0.904 [0.841–0.964] |
| DenseNet-121 | 0.959 [0.923–0.993] | 0.866 [0.816–0.918] | 0.875 [0.786–0.950] |
| EfficientNet-B0 | 0.963 [0.937–0.984] | 0.510 [0.449–0.570] | 0.522 [0.492–0.556] |
| ConvNeXt-Tiny | 0.972 [0.942–0.998] | 0.917 [0.862–0.968] | 0.874 [0.797–0.954] |
| DeiT-Tiny | 0.965 [0.933–0.993] | 0.851 [0.791–0.911] | 0.861 [0.798–0.927] |

Panel B. F1

| Architecture | Baseline | Upper 50% | Lower 50% |
|---|---|---|---|
| ResNet-50 | 0.888 [0.835–0.941] | 0.783 [0.755–0.813] | 0.793 [0.738–0.846] |
| DenseNet-121 | 0.906 [0.865–0.951] | 0.716 [0.693–0.743] | 0.767 [0.692–0.828] |

| Architecture | Baseline | Upper 50% | Lower 50% |
|---|---|---|---|
| EfficientNet-B0 | 0.903 [0.868–0.938] | 0.537 [0.484–0.589] | 0.576 [0.534–0.611] |
| ConvNeXt-Tiny | 0.908 [0.849–0.967] | 0.849 [0.802–0.899] | 0.760 [0.688–0.823] |
| DeiT-Tiny | 0.911 [0.868–0.956] | 0.810 [0.761–0.854] | 0.747 [0.650–0.838] |

Panel C. Mean AUC reduction

| Architecture | Reduction [95% CI] |
|---|---|
| ResNet-50 | 0.063 [0.032–0.092] |
| DenseNet-121 | 0.089 [0.035–0.149] |
| EfficientNet-B0 | 0.447 [0.396–0.495] |
| ConvNeXt-Tiny | 0.076 [0.030–0.130] |
| DeiT-Tiny | 0.109 [0.066–0.157] |

### Figure S5

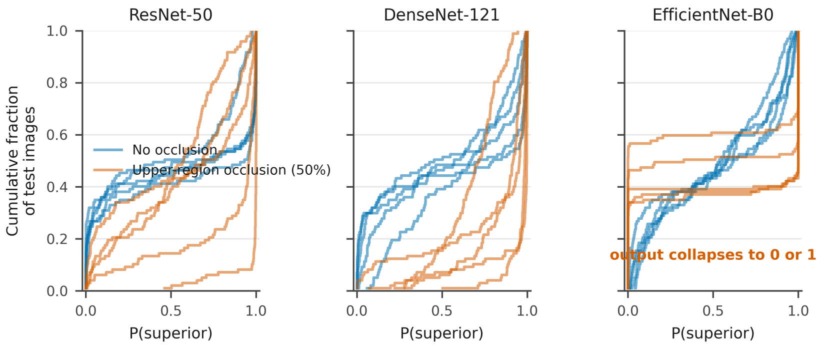


Annotation clarification: "0 or 1" denotes concentration near these limits, not exclusively two exact output values.

**Figure S6**

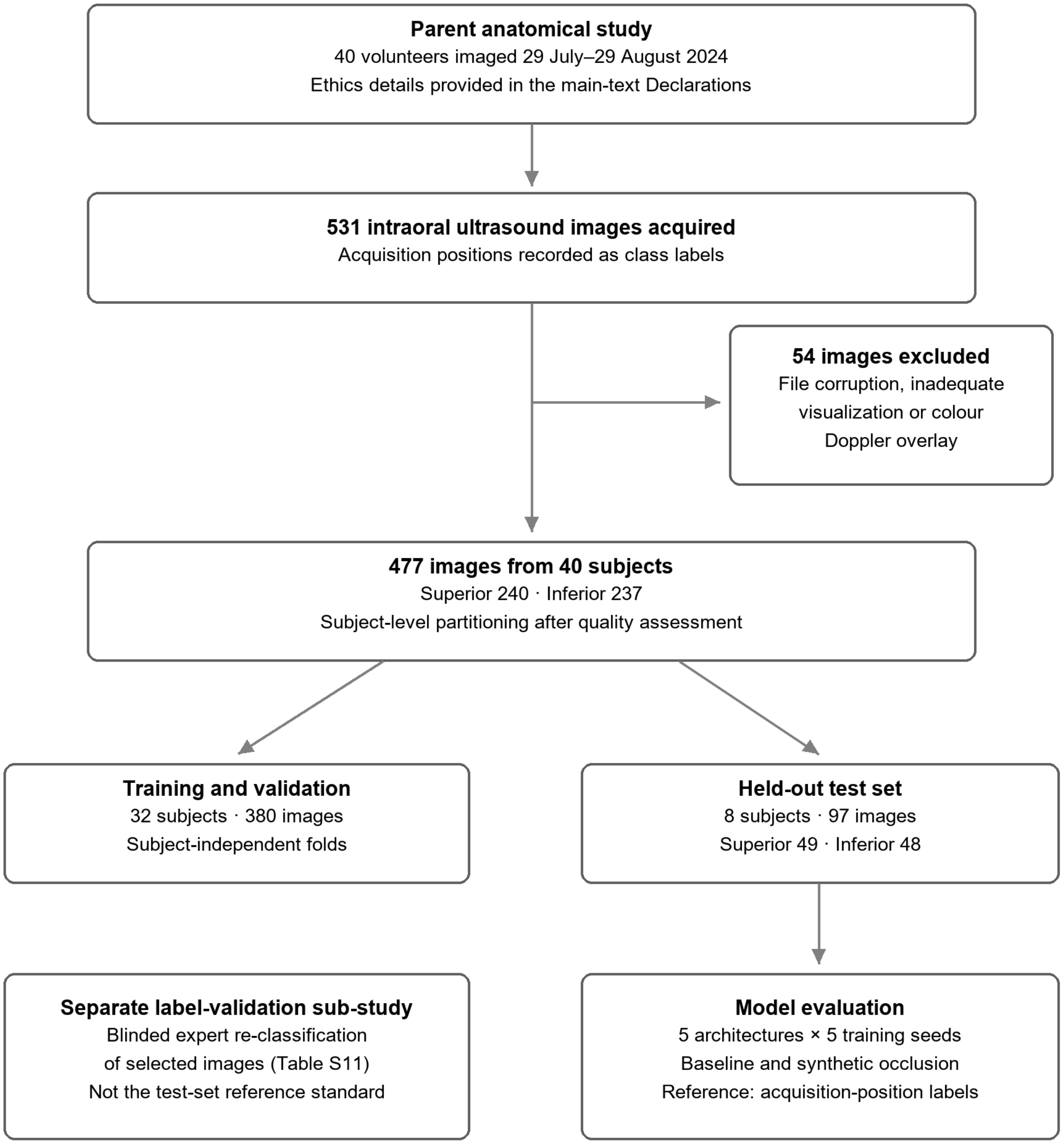

Parent anatomical study
40 volunteers imaged 29 July–29 August 2024
Ethics details provided in the main-text Declarations
531 intraoral ultrasound images acquired
Acquisition positions recorded as class labels
54 images excluded
File corruption, inadequate visualization or colour Doppler overlay
477 images from 40 subjects
Superior 240 · Inferior 237
Subject-level partitioning after quality assessment
Training and validation
32 subjects · 380 images
Subject-independent folds
Held-out test set
8 subjects · 97 images
Superior 49 · Inferior 48
Separate label-validation sub-study
Blinded expert re-classification of selected images (Table S11)
Not the test-set reference standard
Model evaluation
5 architectures × 5 training seeds
Baseline and synthetic occlusion
Reference: acquisition-position labels
